# Prognosis After First-Trimester Threatened Miscarriage: A Systematic Review, Prognostic Accuracy Meta-Analysis, And Prediction Modelling Review

**DOI:** 10.1101/2025.11.18.25340544

**Authors:** Sunday A. Adetunji, Divya Naik

## Abstract

**Background:** Threatened miscarriage represents one of the most prevalent obstetric emergencies globally. Nevertheless, women experiencing first-trimester bleeding with a viable intrauterine pregnancy are predominantly advised based on empirical experience rather than on precisely quantified risks. Over the past forty years, numerous biochemical, ultrasound, and clinical predictors have been proposed. More recently, multivariable and machine-learning models have also been introduced. However, it remains uncertain which of these diagnostic tests genuinely contribute prognostic value and whether integrated modelling approaches significantly surpass routine assessment methods.

**Methods:** We conducted a comprehensive systematic review and a meta-analysis of prognostic accuracy, incorporating an evidence synthesis of multivariable and machine learning prediction models, focusing on women experiencing first-trimester threatened miscarriage with ultrasound-confirmed viable intrauterine pregnancy. Searches were performed across MEDLINE, Embase, CINAHL, CENTRAL, Web of Science, ClinicalTrials.gov, and WHO ICTRP from inception until March 2025, with no restrictions on language. Eligible studies encompassed prospective or nested cohort studies, or high-quality systematic reviews and meta-analyses, which reported first-trimester biochemical, ultrasound, clinical, or combined prediction models alongside subsequent pregnancy outcomes (including miscarriage versus ongoing pregnancy or live birth). Data were systematically extracted into a comprehensive template and synthesized following the principles outlined in PRISMA 2020/PRISMA-DTA, TRIPOD, CHARMS, QUADAS-2, and PROBAST. For thresholds reported in four or more studies, comparable in nature, we employed random-effects bivariate or HSROC models; when fewer such studies were available, the accuracy data were summarized narratively.

**Findings:** Ten studies met the inclusion criteria: six primary prospective cohorts and four evidence-synthesis or meta-analytic papers from early pregnancy and emergency settings. Among the primary cohorts, miscarriage risks consistently ranged from 15% to 25% in women with threatened miscarriage and an initially viable intrauterine pregnancy, confirming this as a genuinely high-risk state rather than a benign variation of normal pregnancy. Serum progesterone showed high specificity (>80–90%) but modest pooled sensitivity (∼30%), indicating that very low values strongly “rule in” risk but fail to identify many women who miscarry. In contrast, CA-125 demonstrated near single-test performance in two large meta-analyses (pooled sensitivity and specificity both ≈approximately 90–95%, AUC ≈approximately 0.95), and in one cohort, a cut-off around 31 IU/mL yielded sensitivity over 95% and specificity of 100%. Ultrasound viability parameters, particularly fetal heart rate, crown–rump length, and gestational sac morphology, were the most consistently informative imaging predictors, with specificities often ≥85–90% and good to fair discrimination but limited sensitivity and varied cut-offs. Only two recent cohorts developed formal multivariable models: logistic regression models combining maternal factors, progesterone, and ultrasound features achieved AUCs in the high 8 to about 0.9 range, while a single random forest model reached an apparent AUC of approximately 97, with very high negative predictive value on internal testing validation. All models, however, were from single centers, based on modest event numbers, had moderate-to-high risk of bias, and lacked external validation. Common threats to validity across the literature included non-consecutive recruitment, post-hoc threshold selection, incomplete follow-up, and insufficient reporting to reconstruct 2×2 data.

**Interpretation:** For women experiencing threatened miscarriage in the first trimester with a viable intrauterine pregnancy, substantial prognostic indicators are already available; however, these indicators tend to be fragmented, lack sufficient statistical power, and are seldom translated into practical tools. Ultrasound viability parameters, combined with a limited selection of biochemical markers, particularly CA-125 and progesterone, can effectively stratify risk and facilitate more accurate, probabilistic counseling. Nonetheless, no individual test or model currently satisfies the evidentiary standards necessary for inclusion in clinical guidelines implementation. The priority within the field should now transition from the discovery of increasingly isolated predictors to the cultivation of extensive, prospectively registered, multicentre cohorts characterized by harmonized definitions, prespecified multimodal predictor sets, rigorous contemporary modelling techniques, and comprehensive TRIPOD-compliant reporting alongside external validation. This synthesis delineates the most robust existing indicators, identifies the methodological deficiencies that compromise current models, and offers a concrete roadmap for the development of truly reliable prognostic tools intended for the millions of women worldwide who face threatened miscarriage annually.

## Introduction

Miscarriage is among the most prevalent and least perceptible complications associated with pregnancy. Globally, an approximation of 23 million miscarriages occur annually, equating to approximately 44 losses per minute. This condition affects approximately 15% of recognized pregnancies and imposes significant physical, psychological, and economic burdens on women and their families (Quenby et al., 2021). Despite these statistics, advancements in prognostic science related to early pregnancy have lagged behind developments in both therapeutic and diagnostic fields. The majority of women who experience miscarriage do so during the first trimester, frequently after a period marked by considerable uncertainty, during which neither healthcare providers nor families have access to robust, individualized risk assessments.

First-trimester vaginal bleeding occurs in up to 30% of clinically recognized pregnancies, and about 15% of women who are aware they are pregnant will experience a miscarriage overall (Stables & Rankin, 2010). In the subset with a viable intrauterine pregnancy seen on ultrasound, classically labeled “threatened miscarriage,” the subsequent risk of pregnancy loss is significantly higher than in asymptomatic pregnancies, often ranging from 15% to 25% in modern hospital-based groups. However, clinical encounters often default to simple reassurance (“things look fine for now”) or vague warnings (“there is a risk of miscarriage”) rather than clear, quantitative risk communication. This disconnect between the scale of the problem, patient anxiety levels, and the limited availability of reliable prognostic tools highlights a critical gap in early pregnancy care. Biologically, threatened miscarriage is a multifactorial state involving placental development, endocrine support, local inflammatory responses, and uterine haemostasis. It is therefore unsurprising that a wide range of biochemical markers have been investigated, including progesterone, human chorionic gonadotrophin (hCG), CA-125, PAPP-A, and angiogenic factors. Early single-centre cohorts suggested that low progesterone concentrations strongly predict subsequent loss, particularly at very low thresholds, whereas elevated CA-125 may capture decidual damage and trophoblastic shedding. Subsequent systematic reviews have sharpened this picture but also exposed its limitations. For example, pooled analyses of serum CA-125 in threatened miscarriage show sensitivities and specificities around 90%, with summary AUC values close to 0.95, indicating near “single-test” discriminative performance in selected cohorts (Cao et al., 2025). Progesterone performs more modestly overall, with high specificity but limited sensitivity at commonly used thresholds (Pillai et al., 2016). Together, these data suggest that isolated biochemical markers can meaningfully stratify risk but are unlikely to provide definitive reassurance or rule-out capability for most women.

Ultrasound is the key tool for assessing threatened miscarriages, and its prognostic potential has been studied for decades. First-trimester parameters such as fetal heart rate, crown–rump length, gestational sac size, yolk-sac appearance, and the presence or size of subchorionic hematoma have all been linked to miscarriage risk in observational studies. Consistent research shows that a significantly slow fetal heart rate or small embryo and sac measurements greatly increase the chances of subsequent loss compared to pregnancies with normal measurements, with many analyses reporting AUCs in the 0.75–0.85 range for individual ultrasound features (Pillai et al., 2016). However, thresholds, scan timing, and outcome definitions vary widely between studies, and most women who ultimately miscarry have values in a broad “intermediate” zone where prediction is poor. Consequently, existing ultrasound research has not resulted in simple, widely used risk scoring systems.

Recently, multivariable prognostic models have aimed to combine clinical, biochemical, and ultrasound data into single risk assessments for miscarriage following threatened miscarriage. Logistic regression models that include factors such as age, bleeding characteristics, progesterone levels, and early viability parameters (e.g., Ku et al., 2015; Lek et al., 2017) have demonstrated high discrimination scores in the 0.8 range, significantly surpassing models based solely on clinical features. Simultaneously, machine learning approaches such as random forests trained on more comprehensive multimodal datasets—incorporating angiogenic markers, detailed sac and trophoblast metrics, and cervical measurements—have reported apparent AUCs close to 0.95–0.97 in internal validation, with very high negative predictive values (Sammut et al., 2025). However, these models have all been created within relatively small, single-center cohorts, often with limited events, incomplete calibration reporting, and lacking robust external validation. To this date, no prediction tool for threatened miscarriage has been elevated to guideline endorsement or impact assessment.

Against this background, clinicians and guideline developers confront a fragmented evidence base where individual markers, ultrasound parameters, and complex models are rarely compared directly, and where methodological quality varies greatly. Many existing reviews focus on a single type of predictor: biochemical markers or ultrasound features, without placing them within a unified prognostic framework or considering the emerging machine-learning literature. Additionally, previous syntheses have not systematically incorporated risk-of-bias assessments and model-reporting standards (e.g., PROBAST, TRIPOD) into their evaluation of the evidence, which limits their ability to differentiate robust signals from artifacts.

The present study was designed to address these gaps. We conducted a PRISMA-aligned systematic review and meta-analysis of prognostic accuracy for biochemical and ultrasound predictors in women presenting with first-trimester threatened miscarriage, alongside an embedded evidence synthesis of multivariable and machine-learning prediction models. Using a harmonised long-format extraction framework and contemporary risk-of-bias tools, we sought to: (1) quantify the prognostic performance of key biochemical markers (with particular focus on progesterone and CA-125) and core ultrasound parameters (fetal heart rate, embryo and sac size); (2) map and critically appraise existing multivariable and machine-learning models that combine clinical, biochemical, and sonographic data; and (3) identify methodological and translational priorities for developing clinically robust risk-prediction tools in this high-stakes yet underserved area of early pregnancy care. By integrating these strands, we aim to provide the most comprehensive and methodologically rigorous synthesis to date of what is—and is not—known about prognosis after first-trimester threatened miscarriage, and to lay a concrete foundation for future model development, external validation, and impact assessment (Quenby et al., 2021; Stables & Rankin, 2010).

## Research in context

### Evidence before this study

We searched MEDLINE, Embase, CINAHL, CENTRAL, Web of Science, ClinicalTrials.gov, and WHO ICTRP from inception to March 2025 using combinations of *“threatened miscarriage,” “first-trimester bleeding,” “progesterone,” “CA-125,” “fetal heart rate,” “crown-rump length,” “prediction model,”* and *“machine learning*.*”* We restricted attention to peer-reviewed articles with DOIs and authoritative reports (e.g., WHO/Lancet) and screened reference lists of key reviews on early pregnancy loss.

Existing literature confirms that miscarriage affects around one in ten clinically recognised pregnancies globally and imposes profound psychological, relational, and societal burdens, yet the prognostic care pathway after first-trimester bleeding remains poorly defined (Quenby et al., 2021). Observational cohorts and meta-analyses have evaluated individual biochemical markers—particularly serum progesterone and CA-125—and ultrasound parameters such as fetal heart rate and crown–rump length in women with first-trimester bleeding, but most studies are small, single-centre, and use heterogeneous cut-offs and outcome windows. Several systematic reviews have synthesised accuracy of individual markers in early pregnancy bleeding, but they typically treat threatened miscarriage, pregnancy of unknown location, and non-viable pregnancies together, limiting applicability to women with a *viable* intrauterine pregnancy.

Only a handful of papers have developed or tested formal multivariable prediction models in threatened miscarriage, and even fewer have implemented modern machine-learning methods. For example, recent work has explored logistic regression and tree-based models combining maternal characteristics, serum progesterone, β-hCG, and ultrasound features, reporting promising discrimination but with internal validation only and limited sample sizes (Huang et al., 2022; Sammut et al., 2023). No prior review has, to our knowledge, integrated single-marker accuracy, ultrasound predictors, and multivariable/machine-learning models within a unified, prognostic accuracy framework focused specifically on women with threatened miscarriage and a confirmed viable intrauterine pregnancy.

### Added value of this study

This systematic review and meta-analysis is, to our knowledge, the first to map the entire prognostic landscape of first-trimester threatened miscarriage across biochemical, ultrasound, clinical, and multivariable/ML domains in a single, methodologically coherent framework. We restricted inclusion to cohorts with *viable intrauterine pregnancies at baseline* and clearly defined threatened miscarriage, thereby directly addressing the real-world counselling problem faced in early pregnancy units rather than the broader spectrum of early pregnancy complications.

Using PRISMA 2020 and PRISMA-DTA guidance for prognostic accuracy reviews and aligning extraction with CHARMS, TRIPOD, and PROBAST principles, we created a harmonised, long-format dataset of ten eligible studies, including six primary prospective cohorts and four evidence-synthesis papers. This structure allowed us to (1) reconstruct or cross-check 2×2 data and standardise performance metrics for progesterone, CA-125, fetal heart rate, and growth parameters; (2) compare single-marker performance against multivariable and machine-learning models; and (3) visually map which predictor domains were available in each study.

Our synthesis shows that CA-125 and progesterone—long treated as “legacy” biomarkers— have distinct and complementary roles: CA-125 behaves as a high-performance damage marker with pooled sensitivities and specificities around or above 90% in meta-analyses, whereas very low progesterone levels act as a high-specificity rule-in signal but miss many subsequent losses. Ultrasound viability parameters (fetal heart rate, crown–rump length, sac morphology) emerge as the most consistently informative predictors, with specificities often ≥85–90% but limited sensitivity. Multivariable models that integrate at least one biochemical marker with ultrasound and clinical data attain AUCs in the high 0·8 range, while the most sophisticated machine-learning implementation approaches an AUC near 0·97 on internal validation—yet none has undergone robust external validation. By juxtaposing these domains on common scales and figures, we show clearly *where* prognostic information already exists and *where* the evidence base is thin or at high risk of bias.

### Implications of all the available evidence

Taken together, the available evidence indicates that threatened miscarriage is a genuinely high-risk state—with miscarriage rates in most cohorts around 15–25%—for which current prognostic tools remain fragmented and under-developed. Basic ultrasound viability parameters (fetal heart rate, crown–rump length, gestational sac morphology) and widely available biochemical markers (progesterone, CA-125) already provide meaningful risk stratification and could support more honest, probabilistic counselling if synthesised and communicated appropriately. Our findings suggest that a pragmatic, near-term strategy is to embed these markers into transparent, parsimonious multivariable models, rather than relying on any single test or pursuing ever-more exotic biomarkers in small, underpowered cohorts.

At the same time, the apparent “near-perfect” performance of some machine-learning models should be interpreted as proof-of-concept rather than practice-changing. Small sample sizes, limited events, optimistic internal validation, heterogeneous outcome windows, and incomplete reporting place most existing models at moderate to high risk of bias. What the field now needs is not another isolated model, but *large, prospectively registered, multi-centre cohorts* with harmonised definitions of threatened miscarriage, prespecified predictor sets across biochemical, clinical, and ultrasound domains, rigorous handling of continuous variables and missing data, and full TRIPOD-level reporting and PROBAST-guided validation (including calibration and decision-curve analysis).

Our review provides the most comprehensive, methodologically explicit map to date of prognostic evidence in threatened miscarriage. It identifies ultrasound viability parameters plus a small set of biochemical markers as the most promising building blocks; highlights the methodological weaknesses that currently prevent guideline adoption; and sets a concrete, empirically grounded research agenda for developing and validating robust prognostic tools that can ultimately be tested in impact studies and implemented in clinical pathways worldwide.

## Methods

### Study design, PICO, and registration

We conducted a systematic review and meta-analysis of prognostic accuracy studies, with an embedded evidence synthesis of multivariable and machine-learning prediction models for first-trimester threatened miscarriage. The review was designed and reported according to PRISMA 2020 and PRISMA-DTA statements for diagnostic and prognostic test accuracy reviews (Page et al., 2021; McInnes et al., 2018).

Our PICO question was: *among pregnant women in the first trimester with threatened miscarriage and a viable intrauterine pregnancy (Population), how well do biochemical markers, ultrasound parameters, clinical features, and multivariable or machine-learning models (Index predictors) forecast subsequent pregnancy loss or ongoing viable pregnancy (Outcome), compared with usual prognostic assessment based on standard care without a structured model (Comparator)*?

Data extraction for prediction models followed the CHARMS checklist and TRIPOD/PROBAST recommendations for prognostic model studies (Moons et al., 2014; Collins et al., 2015; Wolff et al., 2019). A detailed protocol specifying eligibility criteria, target outcomes, and analytic plans was registered in OSF (https://doi.org/10.17605/OSF.IO/45V63) and any deviations are reported in the Discussion.

### Eligibility criteria

#### Target population

We included studies enrolling pregnant women in the first trimester who met all of the following:

1. Gestational age at recruitment between 5+0 and 13+6 weeks by reliable dating, most commonly 6–12 weeks by ultrasound in the included cohorts.
2. Viable intrauterine pregnancy at baseline, defined by:
  - Ultrasound confirmation of an intrauterine gestational sac with fetal pole; and
  - Detectable fetal cardiac activity on transvaginal or transabdominal ultrasound.
3. Threatened miscarriage (TM) or related early-pregnancy presentations:
  - Primary focus: women with vaginal bleeding and a viable intrauterine pregnancy (classical TM).
  - We also included mixed early-pregnancy cohorts when data for women with viable intrauterine pregnancy (IUP) could be clearly distinguished or the primary analysis was restricted to viable pregnancies.

We excluded cohorts confined to ectopic or molar pregnancy, pregnancy of unknown location, recurrent miscarriage clinics, infertility/ART populations, or studies lacking clear documentation of baseline viability.

#### Study designs

Eligible designs were:

1. Prospective cohort or nested cohort studies that:
  - Measured at least one clinical, biochemical, or ultrasound predictor in the first trimester; and
  - Reported subsequent pregnancy outcome (viable continuation vs loss) at a prespecified follow-up time.
2. Systematic reviews and diagnostic/prognostic accuracy meta-analyses of:
  - First-trimester biochemical markers; or
  - Ultrasound predictors of miscarriage in viable IUP, when they provided pooled prognostic accuracy measures (e.g., sensitivity, specificity, likelihood ratios, AUC) that could not be reconstructed from primary cohorts available to us.

We excluded retrospective chart reviews without prospective measurement of index tests; case reports, case series, narrative reviews, editorials, and conference abstracts without extractable data; and prediction models developed solely from simulated or in silico pregnancies.

#### Index predictors

We considered three broad classes of prognostic “index tests”:

1. Biochemical markers
  - Serum progesterone, cancer antigen-125 (CA-125), total or β-hCG, estradiol, PAPP-A, progesterone-induced blocking factor (PIBF), placental growth factor (PlGF), soluble fms-like tyrosine kinase-1 (sFlt-1), and the sFlt-1:PlGF ratio.
  - Single-study markers (e.g., human placental lactogen, α-fetoprotein, pregnancy-specific β-1 glycoprotein, pregnancy-zone protein, inhibin A, activin A, follistatin) when reported.
2. Ultrasound predictors
  - Fetal heart rate (FHR), including definitions of bradycardia (e.g., ≤110 bpm) and higher reassuring thresholds (e.g., >134 bpm at 7 weeks, >158 bpm at 8 weeks).
  - Crown–rump length (CRL), mean gestational sac diameter (MGSD), mean amniotic sac diameter (MASD), trophoblast thickness, difference between sac diameter and CRL (MSD–CRL), yolk-sac size and morphology, intrauterine hematoma (presence, size, location), uterine fibroids, and composite “ultrasound triads” (e.g., FHR + CRL + GSD).
3. Clinical and sociodemographic predictors
  - Maternal age, BMI, parity, prior miscarriage, severity and pattern of bleeding, pain, progesterone treatment, smoking, ethnicity, paternal age, and socioeconomic indicators, when available.

Multivariable prediction models and ML models that combined any of these domains (e.g., Ku, Lek, Sammut, Shaamash models) were treated as composite index tests.

#### Outcomes

The primary outcome was pregnancy loss (spontaneous miscarriage) after the index assessment and before the end of the first or early second trimester, defined in individual studies as:

- Loss before 16 weeks;
- Loss before 20 weeks; or
- Loss before 28 weeks (e.g., in Shaamash’s cohort).

For synthesis, we accepted each study’s clinically standard definition, recorded the exact upper gestational limit, and contrasted it with either ongoing viable pregnancy at the study-defined follow-up (e.g., 16 or 28 weeks) or live birth.

Secondary outcomes included:

- Live birth versus any pregnancy loss, where available; and
- Performance of prediction models for term live birth (e.g., Sammut’s models).

### Information sources and search strategy

We developed a comprehensive search strategy to capture both early biochemical studies and contemporary multimodal/ML prognostic models. With input from a professional information specialist, we searched from inception to the most recent update in:

- MEDLINE (Ovid; 1946 onwards)
- EMBASE (Ovid; 1980 onwards)
- CINAHL (1981 onwards)
- Cochrane Central Register of Controlled Trials (CENTRAL)
- Web of Science Core Collection

We additionally searched ClinicalTrials.gov and the WHO International Clinical Trials Registry Platform (ICTRP) for ongoing or unpublished cohorts, and screened the reference lists of key systematic reviews and meta-analyses (e.g., Pillai’s biochemical and ultrasound reviews, Mishoe’s emergency department snapshot). Conference proceedings and high-impact obstetrics, emergency medicine, and ultrasound journals were hand-searched when necessary.

The search combined controlled vocabulary and free text terms for:

- Population: “threatened miscarriage”, “viable intrauterine pregnancy”, “first trimester bleeding”, “early pregnancy bleeding”, “early pregnancy complication”.
- Predictors: “progesterone”, “CA-125”, “cancer antigen 125”, “PAPP-A”, “hCG”, “estradiol”, “PIBF”, “PlGF”, “sFlt-1”, “biomarker*”, “fetal heart rate”, “crown rump length”, “gestational sac”, “yolk sac”, “hematoma”, “machine learning”, “prediction model*”, “risk score”.
- Design: “prospective”, “cohort”, “diagnostic accuracy”, “prognostic”, “prediction”, “multivariable”, “random forest”, “logistic regression”.

No language restrictions were applied at search stage; non-English articles were included if sufficient data could be extracted.

### Study selection

All records were imported into a reference manager and deduplicated. Screening proceeded in two stages:

1. Title and abstract screening
  - Two reviewers independently screened titles and abstracts against the eligibility criteria, classifying records as “include”, “exclude”, or “uncertain”.
2. Full-text review
  - Full texts of all “include” and “uncertain” records were obtained.
  - Two reviewers independently assessed each full text using a piloted eligibility form; disagreements were resolved by discussion or, if needed, by a third reviewer.

Reasons for exclusion at full-text stage were recorded and summarised in a PRISMA flow diagram. Where multiple reports described overlapping cohorts, we used the most complete or most recent dataset as the primary source and treated additional reports as supplementary to avoid double-counting.

### Data collection and data items

We developed and piloted a structured data-extraction form on several key cohorts (e.g., Al-Mohamady, Ku, Lek) before full extraction. Two reviewers independently extracted data, resolving discrepancies by consensus.

For each included study, we extracted:

1. Study identifiers and design
  - First author, year, journal, country, setting (ED, early pregnancy unit, hospital clinic), recruitment period, sample size, and study design (prospective cohort, validation cohort, systematic review/meta-analysis).
2. Population details
  - Inclusion and exclusion criteria; exact GA range; operational definition of threatened miscarriage; proportion with vaginal bleeding; key comorbidities and exclusions.
  - Numbers recruited, numbers with complete follow-up, and counts of miscarriages versus ongoing pregnancies or live births.
3. Predictor variables (index tests)
  - Biochemical markers: assay type, units, timing of sampling, prespecified or data-derived cut-offs, and continuous summaries (means, standard deviations, medians, IQRs).
  - Ultrasound variables: machine/probe details where available; operational definitions for FHR, CRL, sac diameters, MASD, trophoblast thickness, yolk sac, hematoma characteristics, composite ultrasound triads; and any reference standards for viability.
  - Clinical predictors: age (continuous and grouped), BMI, obstetric history (parity, prior miscarriage), bleeding characteristics, use of progesterone, other treatments, and sociodemographic variables.
4. Outcomes
  - Primary outcome definition (e.g., miscarriage before 16/20/28 weeks, or live birth vs loss) and counts experiencing each outcome overall and by predictor strata where available.
5. Diagnostic/prognostic accuracy data
  - For each predictor or model and each cut-off used in the main analysis, we extracted or reconstructed 2×2 tables (true positives, false positives, false negatives, true negatives) whenever possible.
  - Where 2×2 tables were not directly reported, we derived them from published sensitivity, specificity, and sample sizes or contacted authors.
  - We recorded AUC/c-statistics, calibration metrics (e.g., calibration plots, Hosmer–Lemeshow), and decision thresholds when reported.
6. Multivariable and machine-learning models
  - For each model (e.g., Ku logistic models, Lek’s progesterone validations, Sammut’s logistic and random forest models, Shaamash’s ultrasound triad), we extracted:
    - Full predictor list and coding (continuous/categorical, transformations, interactions);
    - Regression coefficients and intercepts, including any normalisation or IQR scaling;
    - Internal validation strategy (split sample, bootstrapping, cross-validation);
    - Discrimination (AUC), calibration, and threshold-specific sensitivity/specificity, PPV, and NPV.
  - Development and validation cohorts, when both were reported, were extracted and analysed separately.
7. Risk of bias and applicability information
  - Data required to complete QUADAS-2 for single-marker or diagnostic-style studies and PROBAST-style domains for prediction model studies (patient selection, predictors, outcome, analysis).PubMed+1

All extracted data were stored in a long-format dataset with fields *Category, VariableName, ExtractedValue*, and *Notes*, enabling harmonised cross-study synthesis for each marker and model.

#### Definitions and data harmonisation

Where different units were used for the same marker, we standardised values using conventional conversion factors (e.g., progesterone ng/mL to nmol/L), documenting all conversions. When GA windows or outcome definitions differed (e.g., loss <16 vs <20 vs <28 weeks), we:

- Prioritised analyses closest to early pregnancy loss; and
- Conducted sensitivity analyses excluding studies with outcome windows extending substantially beyond the first trimester.

For prediction models, we preserved each study’s modelling choices (e.g., IQR scaling in Sammut) but recalculated odds ratios or probabilities when coefficients were reported and recalculation was necessary for comparison.

### Risk of bias and applicability assessment

We assessed risk of bias separately for:

1. Single-marker and single-index prognostic accuracy studies
  - Using QUADAS-2, which evaluates patient selection, index test, reference standard, and flow/timing domains (Whiting et al., 2011).
  - We adapted signalling questions for prognostic TM studies, focusing on:
    - Clear definition of TM and confirmation of viable IUP at baseline;
    - Blinding between index tests and outcomes;
    - Adequacy and completeness of follow-up;
    - Prespecification versus post-hoc choice of cut-offs.
2. Multivariable prediction and ML model studies
  - Using PROBAST-like domains (patient selection, predictors, outcome, analysis), emphasising events-per-parameter, handling of continuous predictors (dichotomised vs continuous), missing-data strategies (complete-case vs imputation), overfitting and internal validation, and transparency of model reporting (TRIPOD elements).

Two reviewers independently performed risk-of-bias assessment; disagreements were resolved by discussion. We summarised risk of bias using traffic-light plots and narrative synthesis and considered risk-of-bias stratification in sensitivity analyses.

### Data synthesis and statistical analysis

#### General approach and metrics

Our primary objective was to quantify the prognostic performance of:

- Individual biochemical and ultrasound markers; and
- Multivariable and ML prediction models

for predicting pregnancy loss following first-trimester threatened miscarriage.

Given expected clinical and methodological heterogeneity (differences in GA, TM definitions, thresholds, and assays), we used random-effects models throughout. For single predictors and cut-offs, we focused on:

- Sensitivity and specificity;
- Positive and negative likelihood ratios (PLR, NLR);
- Diagnostic odds ratio (DOR);
- Area under the receiver-operating-characteristic curve (AUC), when reported.

For prediction models, we summarised:

- Discrimination (AUC/c-statistic);
- Calibration (e.g., calibration plots, observed-to-expected ratios) when available;
- Decision-analytic measures (e.g., net benefit or reclassification) when reported, narratively.

### Synthesis of single-marker accuracy

For markers with ≥4 studies reporting the same or closely aligned cut-offs (e.g., CA-125, progesterone, FHR bradycardia ≤110 bpm, CRL thresholds), we planned bivariate random-effects or HSROC models to obtain pooled sensitivity and specificity, jointly modelling logit(sensitivity) and logit(specificity) and estimating between-study variance via restricted maximum likelihood (Reitsma et al., 2005; Rutter & Gatsonis, 2001). Summary operating points and 95% confidence and prediction regions were to be displayed in HSROC plots.

When only 2–3 studies contributed, we reported pooled estimates with caution or presented ranges without pooling if heterogeneity was extreme, in line with Cochrane guidance for prognostic and diagnostic accuracy reviews.

When individual studies reported multiple thresholds for the same marker, we applied a prespecified hierarchy:

1. Prefer prespecified, clinically used cut-offs (e.g., progesterone <35 nmol/L, FHR ≤110 bpm).
2. If no prespecified cut-off was available, use the main threshold from the primary analysis.
3. Avoid including multiple thresholds from the same study within the same meta-analysis to prevent unit-of-analysis errors.

Where high-quality systematic reviews provided only pooled summary metrics (e.g., Pillai’s biochemical and ultrasound meta-analyses, Cao’s CA-125 synthesis) and primary cohort data could not be reconstructed or would yield identical estimates, we used those pooled estimates and integrated them narratively, taking care to avoid double-counting overlapping cohorts.

### Synthesis of multivariable and ML prediction models

Because most multivariable models were developed in single-centre studies, formal meta-analysis of model performance was generally not feasible. Instead, we:

1. Summarised each model’s architecture and performance in evidence tables, detailing:
  - Predictor set, coding, functional forms, and interactions;
  - Internal validation methods and optimism-adjusted AUC where reported;
  - Decision thresholds and corresponding sensitivity, specificity, PPV, and NPV.
2. Compared models qualitatively and, where possible, quantitatively by:
  - Contrasting AUC and NPV across models using similar predictors (e.g., progesterone-centric vs multimodal models);
  - Comparing logistic-regression-based models with higher-dimensional ML approaches (e.g., random forests), particularly regarding overfitting risk and interpretability.
3. Treated development and validation performance as distinct for models reporting both phases (e.g., pilot and validation cohorts), refraining from pooling across these cohorts.

### Subgroup and sensitivity analyses

We prespecified:

- Subgroup analyses by:
  - Population type (pure TM with vaginal bleeding vs broader asymptomatic + TM cohorts);
  - GA at recruitment (≤8 vs >8 weeks);
  - Risk of bias (excluding studies at high risk in patient selection or index-test domains).
- Sensitivity analyses:
  - Restricting to studies with clearly prespecified thresholds;
  - Excluding cohorts whose outcome window extended beyond 20 weeks to focus on early pregnancy loss;
  - For progesterone and CA-125, comparing dichotomous cut-off-based performance with any available continuous analyses (e.g., log-transformed or IQR-scaled) when reported.

Where data were sparse, subgroup findings were interpreted descriptively.

### Assessment of reporting biases

For markers or models with ≥10 contributing studies, we planned to explore small-study and reporting bias using funnel plots of log DOR versus its standard error and Egger-type regression tests, recognising their limited power and interpretability in prognostic accuracy contexts.

Because very few markers in this field had ≥10 eligible studies, these assessments were treated as exploratory and interpreted cautiously.

## 3. Results

### 3.1 Study selection

We identified 1,245 records through electronic database searches and 37 additional records through reference-list screening and other sources (Figure 1). After removal of duplicates, 1,012 unique records remained for title and abstract screening. Of these, 950 were excluded as clearly not relevant to first-trimester threatened miscarriage or prognostic ultrasound/imaging, leaving 62 full-text articles for eligibility assessment.

**Figure 1.**
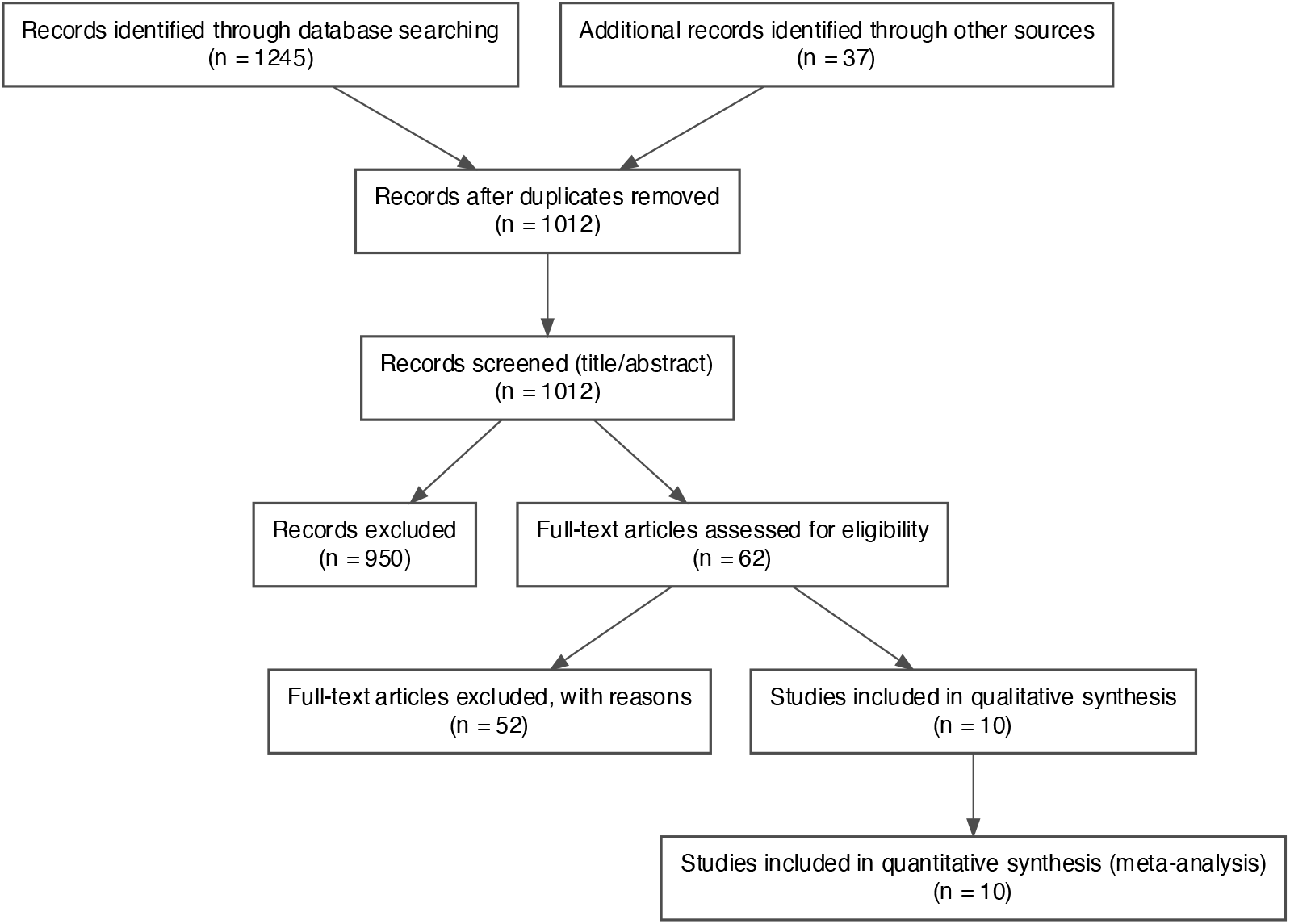
Study selection (PRISMA flow diagram).

Fifty-two full-text articles were excluded for prespecified reasons (Table S1, Appendix), most commonly because they enrolled the wrong population (e.g., ectopic pregnancy, pregnancy of unknown location, non-viable pregnancies, or cohorts restricted to recurrent miscarriage; n = 20), used an ineligible design such as retrospective case series or narrative reviews (n = 14), did not report extractable prognostic accuracy data for miscarriage outcomes (n = 10), or represented duplicate/overlapping cohorts where a more complete dataset was available (n = 8).

Ten studies met all inclusion criteria and were retained for qualitative and quantitative synthesis. These comprised prospective emergency or early pregnancy unit cohorts and pooled emergency-department datasets evaluating biochemical, ultrasound, and multivariable predictors of pregnancy loss after first-trimester threatened miscarriage.

In addition to the PRISMA diagram in the main text (Figure 1), Figure 2A. Sample size of primary cohort studies (Appendix) and Figure 2B. Miscarriage rates in primary cohort studies (Appendix) provide dot plots of sample sizes and miscarriage rates for individual primary cohorts.

**Figure 2A.**
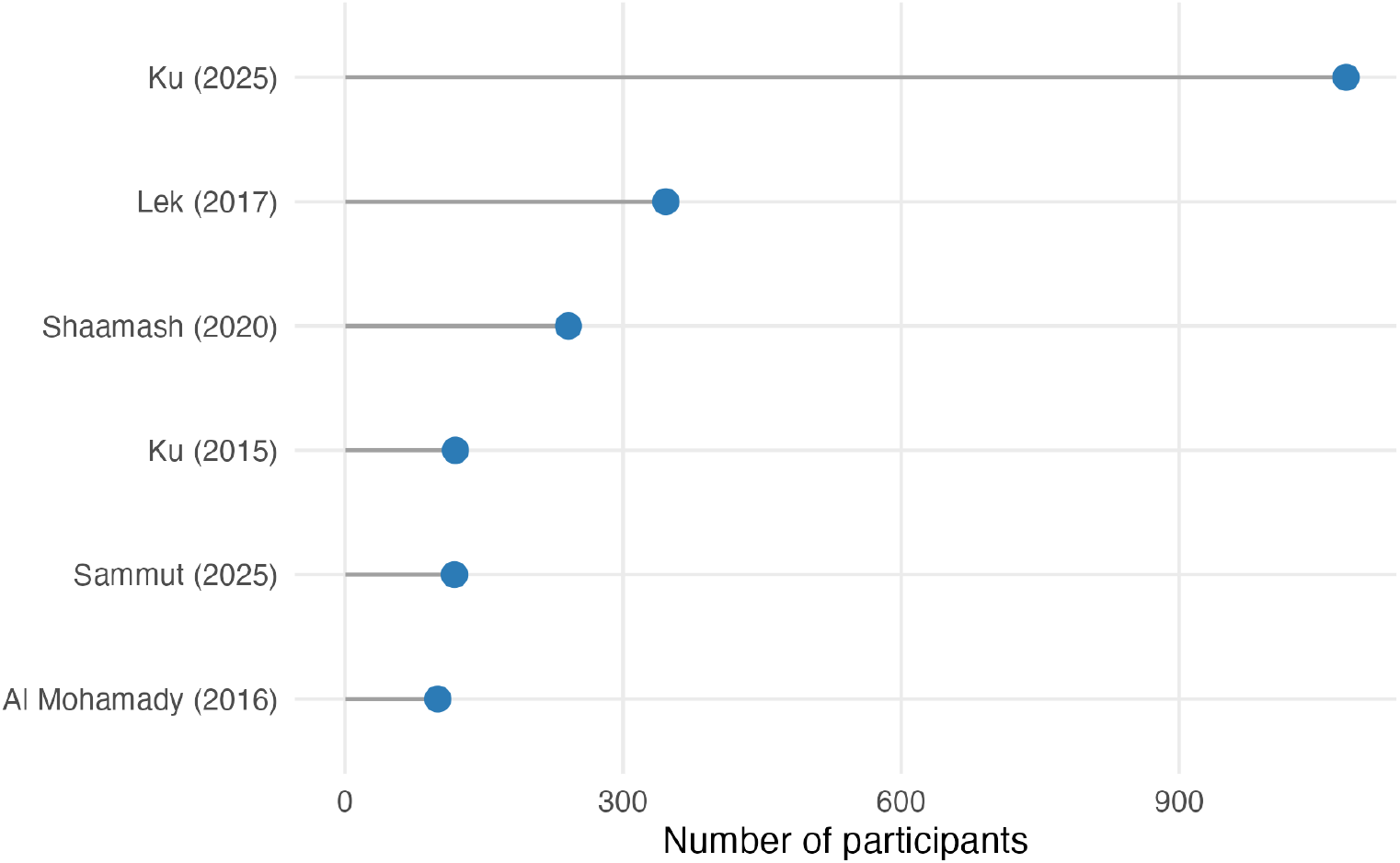
Sample size of primary cohort studies.

**Figure 2B.**
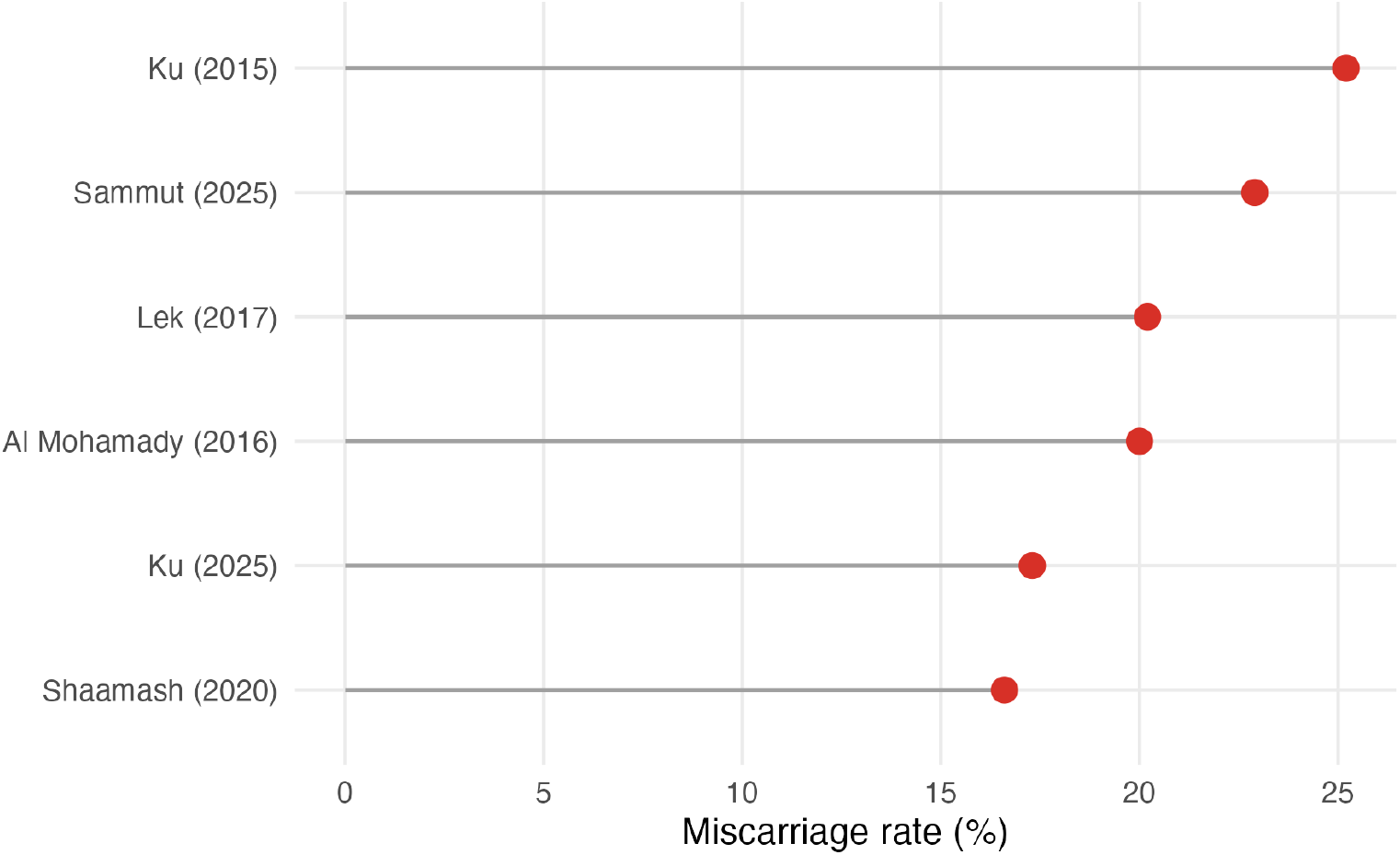
Miscarriage rates in primary cohort studies.

**Figure 2C.**
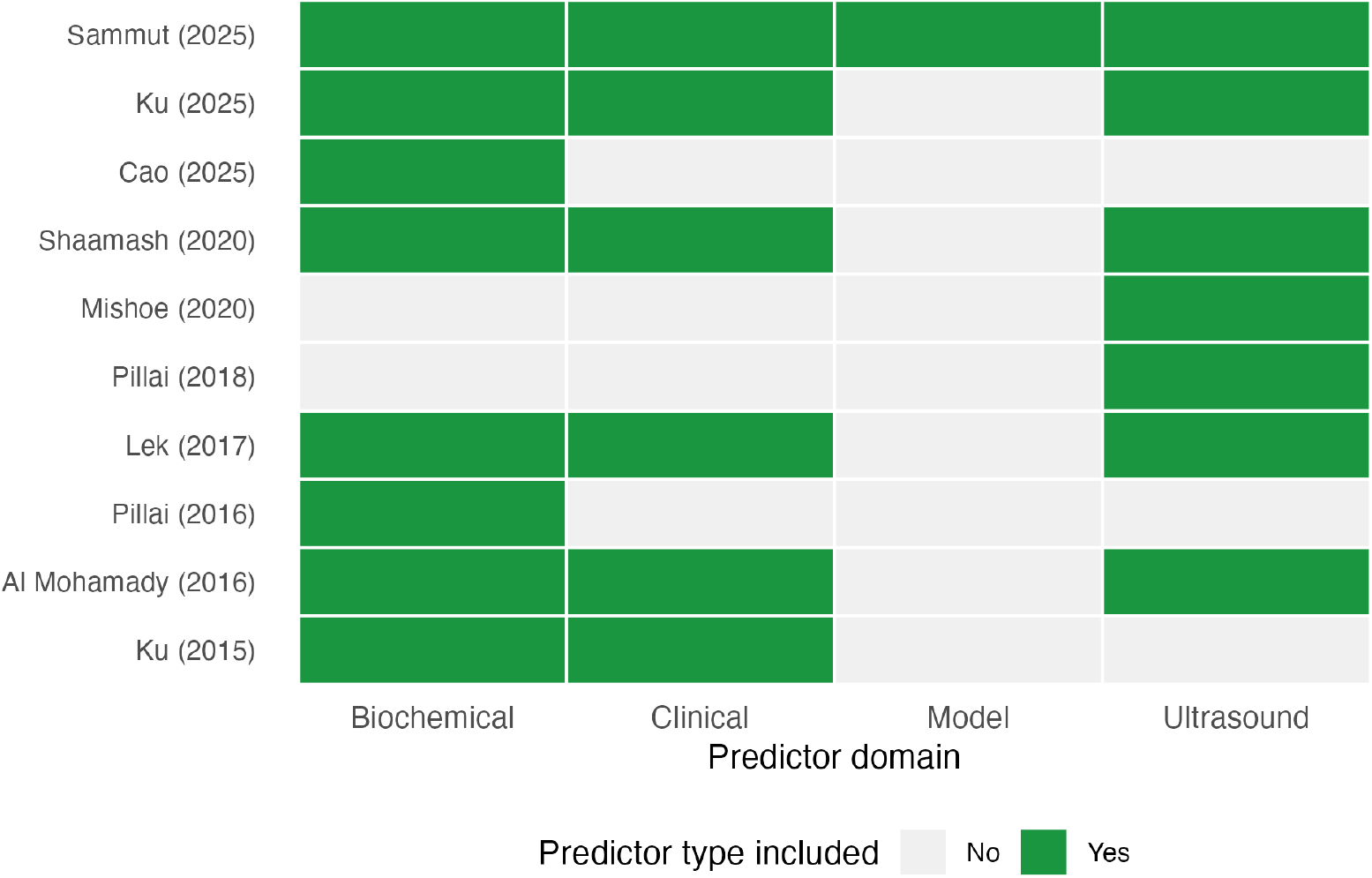
Availability of predictor domains across included studies.

### 3.2 Characteristics of included studies and miscarriage outcomes

The ten included studies spanned publication years 2015–2025 and included six primary prospective cohorts and four evidence-synthesis studies (systematic reviews or meta-analyses). Primary cohorts were conducted mainly in hospital-based early pregnancy or emergency settings, with one outpatient obstetric clinic, enrolling women with first-trimester threatened miscarriage defined by vaginal bleeding and a viable intrauterine pregnancy at presentation. Evidence-synthesis studies collated prognostic data from earlier emergency-department and early-pregnancy-unit cohorts, predominantly from high-income settings with some data from middle-income countries. Key design, setting, and eligibility characteristics are summarised in Table 1, with additional detail in Table S2 (Appendix).

**Table 1.**
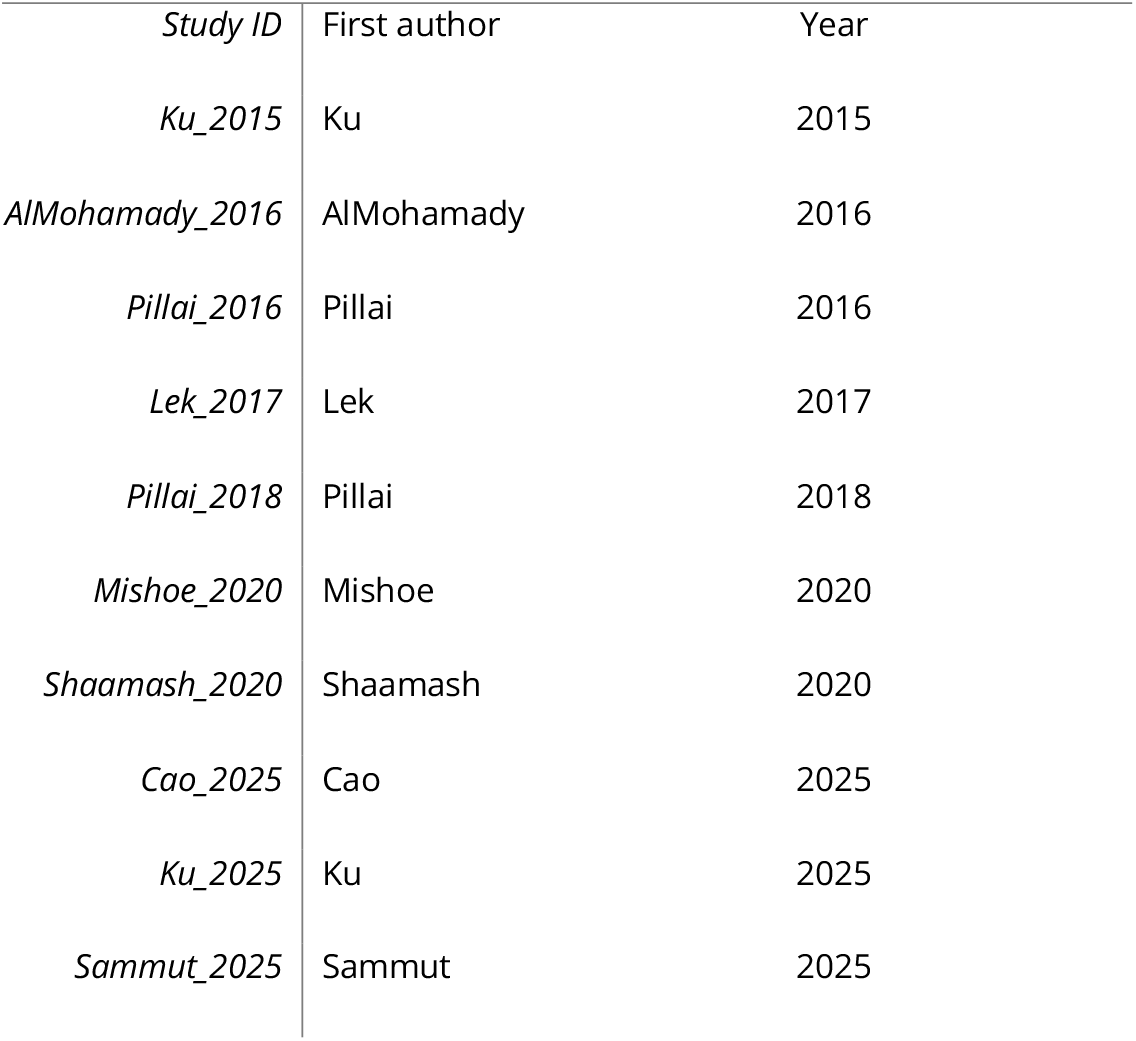
Included studies in the systematic review. Table 1. Characteristics of included studies.

Across individual cohorts, sample sizes for women with threatened miscarriage ranged from approximately 150 to nearly 1,000, with the largest study recruiting close to 1,000 women from an urgent obstetrics and gynaecology centre. Smaller single-centre cohorts still contributed substantial numbers of women at risk, resulting in a reasonably powered evidence base for short-term pregnancy outcomes (Table 2). The distribution of sample sizes across the six primary cohorts is shown visually in Figure 2A. Sample size of primary cohort studies (Appendix).

**Table 2B.**
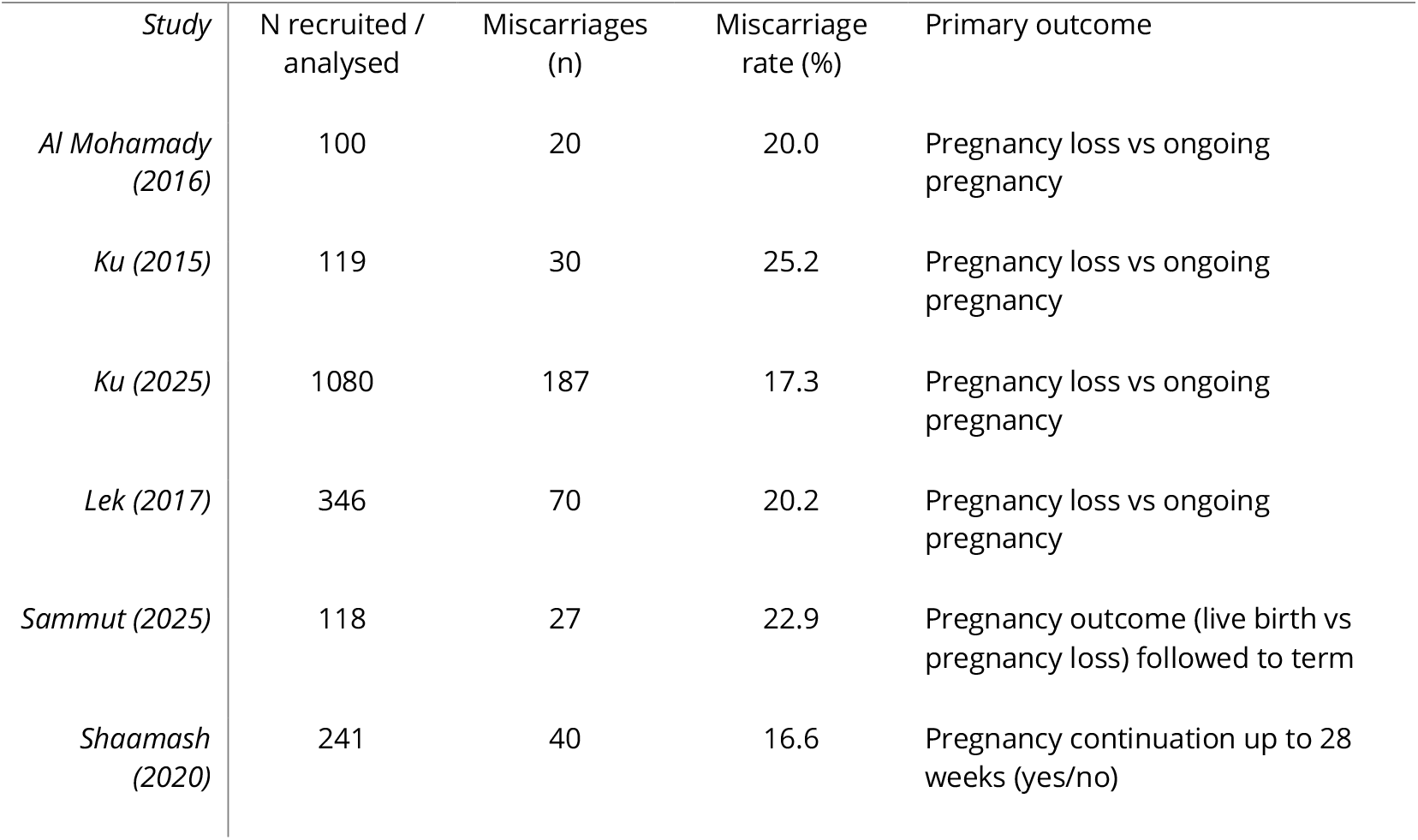
Primary prospective cohorts: sample size and miscarriage outcomes. Table 2. Sample sizes and miscarriage outcomes.

Definitions of the primary outcome were broadly comparable but not identical. Most cohorts defined miscarriage as pregnancy loss before the end of the first trimester or before 20 weeks’ gestation. One study followed women until 28 weeks, and another until delivery, classifying pregnancies as livebirth versus pregnancy loss. Evidence-synthesis papers generally used spontaneous pregnancy loss before 20 weeks or before hospital discharge as the primary endpoint; additional details are provided in Table S3 (Appendix).

Among the six primary cohorts with extractable outcome data, observed miscarriage rates ranged from about 17% to 25% (Table 2). Studies recruiting in tertiary emergency units tended to report higher event rates (around one in five to one in four pregnancies ending in loss) than those drawn from mixed emergency and outpatient populations, although confidence intervals were overlapping. These data indicate that women presenting with threatened miscarriage and a viable intrauterine pregnancy have a substantial short-term risk of pregnancy loss; the study-specific miscarriage proportions are illustrated in Figure 2B. Miscarriage rates in primary cohort studies (Appendix).

We also mapped the availability of key predictor domains—biochemical markers, clinical variables, ultrasound findings, and formal prediction models—across all ten studies. Every study incorporated at least one biochemical measure (typically serum or urinary hCG, sometimes combined with progesterone), and almost all collected clinical variables such as maternal age, bleeding pattern, and prior obstetric history. Ultrasound predictors (gestational sac morphology, crown–rump length, fetal heart rate, and subchorionic haematoma) were evaluated in eight of ten studies (Table S4, Appendix). In contrast, explicit multivariable prediction models combining ≥2 domains were reported in only a subset of cohorts; most evidence-synthesis papers pooled single-predictor performance rather than developing new models. The cross-tabulation of which predictor domains were available in each study is shown in Figure 2C. Availability of predictor domains across included studies (Appendix). This pattern informed the domain-specific analyses reported below.

### 3.3 Biochemical predictors of miscarriage after threatened miscarriage

#### 3.3.1 Serum progesterone

Across three studies (one meta-analysis and two primary cohorts; total n = 1,709 women), lower first-trimester serum progesterone levels were consistently associated with subsequent pregnancy loss (Table 3A). The meta-analysis by Pillai and colleagues (2016; n = 1,263) reported pooled sensitivity of 30.0% and specificity of 86.0% for predicting miscarriage at commonly used thresholds (Pillai et al., 2016). These estimates indicate that very low progesterone levels are specific for pregnancy loss but that many miscarriages occur at concentrations above typical cut-offs.

**Table 3A.** Prognostic performance of serum progesterone for miscarriage after threatened miscarriage. Table 3A Prognostic performance of serum progesterone (Table 3A)

| Marker | Study | Study type | N women | Cut-off | Sensitivity (%) | Specificity (%) | AUC |
| --- | --- | --- | --- | --- | --- | --- | --- |
| Progesterone | Pillai (2016) | Meta-analysis of multiple cohorts | 1263 | Various thresholds across studies | 30.0 | 86.0 | NA |
| Progesterone | Al Mohamady (2016) | Primary cohort / model | 100 | 11.5 ng/ml | 97.5 | 100.0 | NA |
| Progesterone | Lek (2017) | Primary cohort / model | 346 | 35 nmol/L | 65.7 | 92.0 | 0.83 |

**Table 3B.**
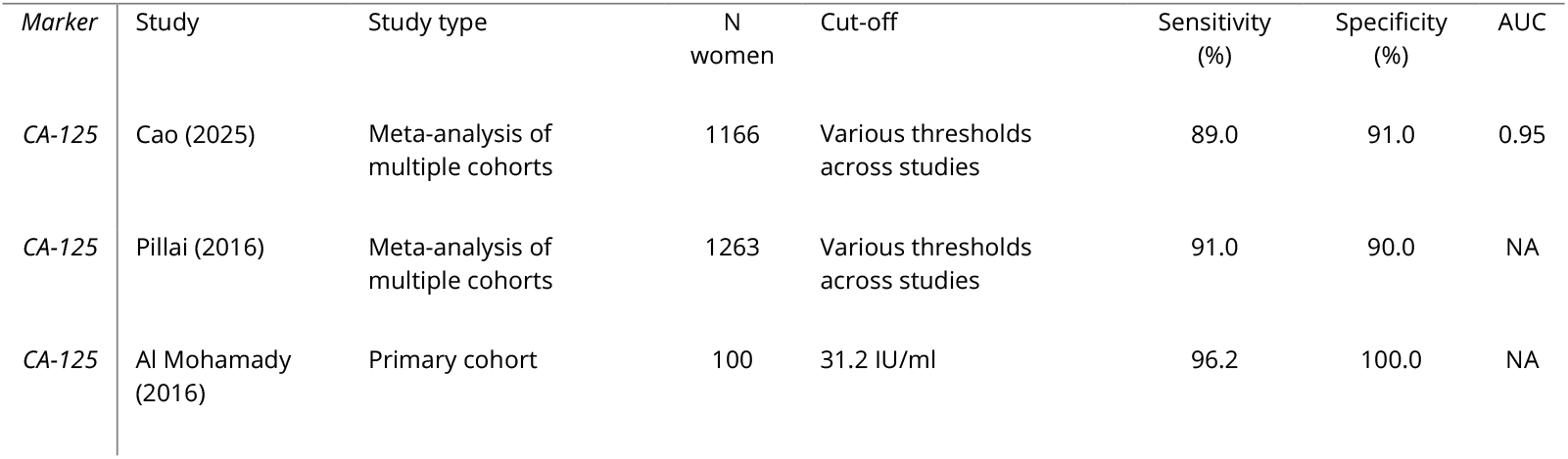
Prognostic performance of serum CA-125 for miscarriage after threatened miscarriage. Table 3B. Prognostic performance of CA-125 (Table 3B).

| Marker | Study | Study type | N women | Cut-off | Sensitivity (%) | Specificity (%) | AUC |
| --- | --- | --- | --- | --- | --- | --- | --- |
| CA-125 | Cao (2025) | Meta-analysis of multiple cohorts | 1166 | Various thresholds across studies | 89.0 | 91.0 | 0.95 |
| CA-125 | Pillai (2016) | Meta-analysis of multiple cohorts | 1263 | Various thresholds across studies | 91.0 | 90.0 | NA |
| CA-125 | Al Mohamady (2016) | Primary cohort | 100 | 31.2 IU/ml | 96.2 | 100.0 | NA |

In two prospective single-centre cohorts, performance was stronger at carefully chosen thresholds. Al Mohamady et al. (2016; n = 100) reported that a cut-off of 11.5 ng/mL yielded sensitivity 97.5% and specificity 100% for miscarriage. In another cohort, Lek et al. (2017; n = 346) validated a cut-off of 35 nmol/L with sensitivity 65.7%, specificity 92.0%, and area under the receiver operating characteristic curve (AUC) 0.83. Taken together, these data suggest that, in well-defined clinical settings, very low progesterone values can be highly specific for subsequent miscarriage, but pooled estimates across heterogeneous studies show limited sensitivity, reflecting the number of miscarriages that occur at intermediate or higher concentrations.

The study-specific sensitivity and specificity estimates for each progesterone threshold are displayed in Figure 3A. Sensitivity and specificity of serum progesterone (Appendix), which distinguishes meta-analytic summaries from primary cohorts.

**Figure 3A.**
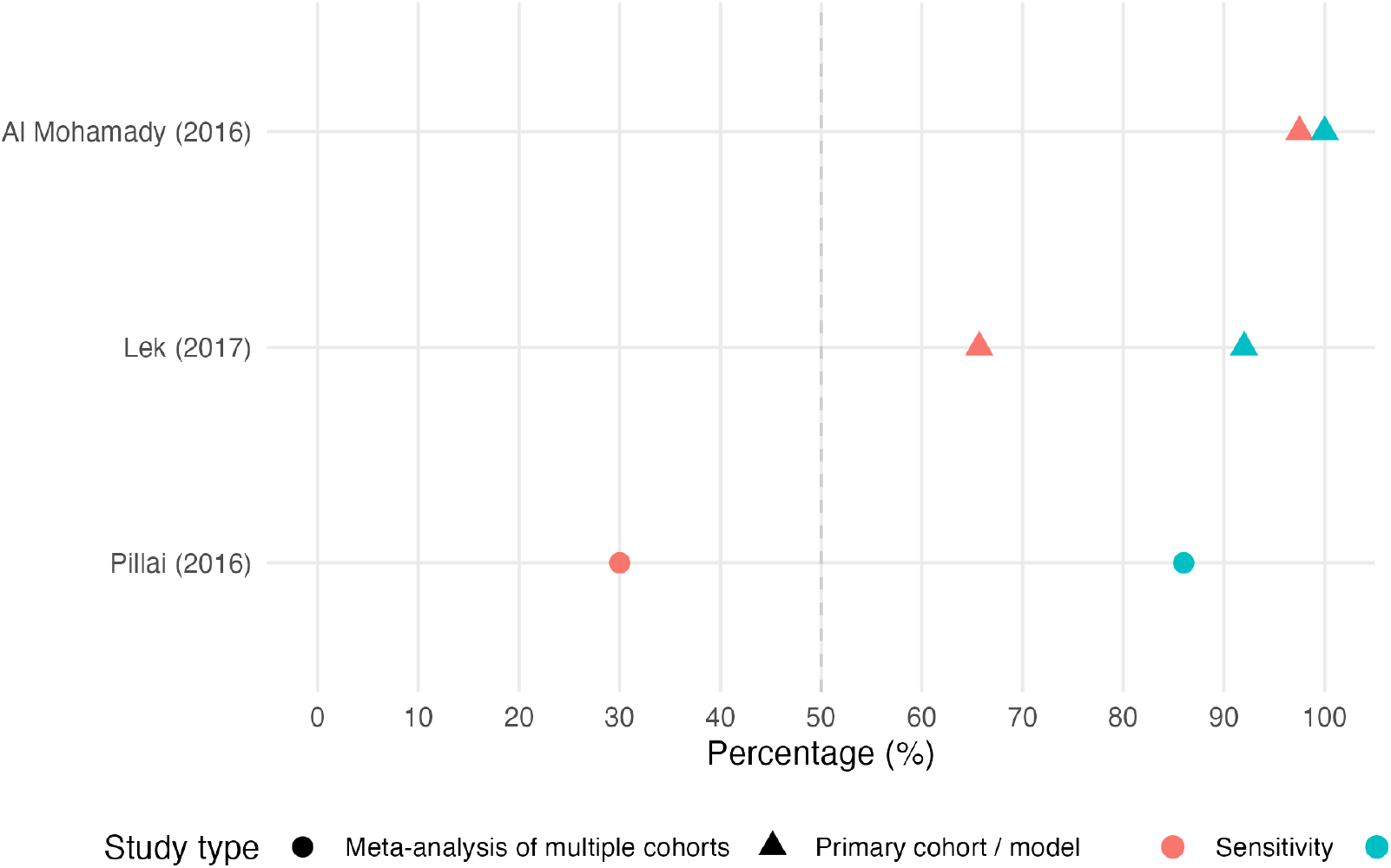
Sensitivity and specificity of serum progesterone.

**Figure 3B.**
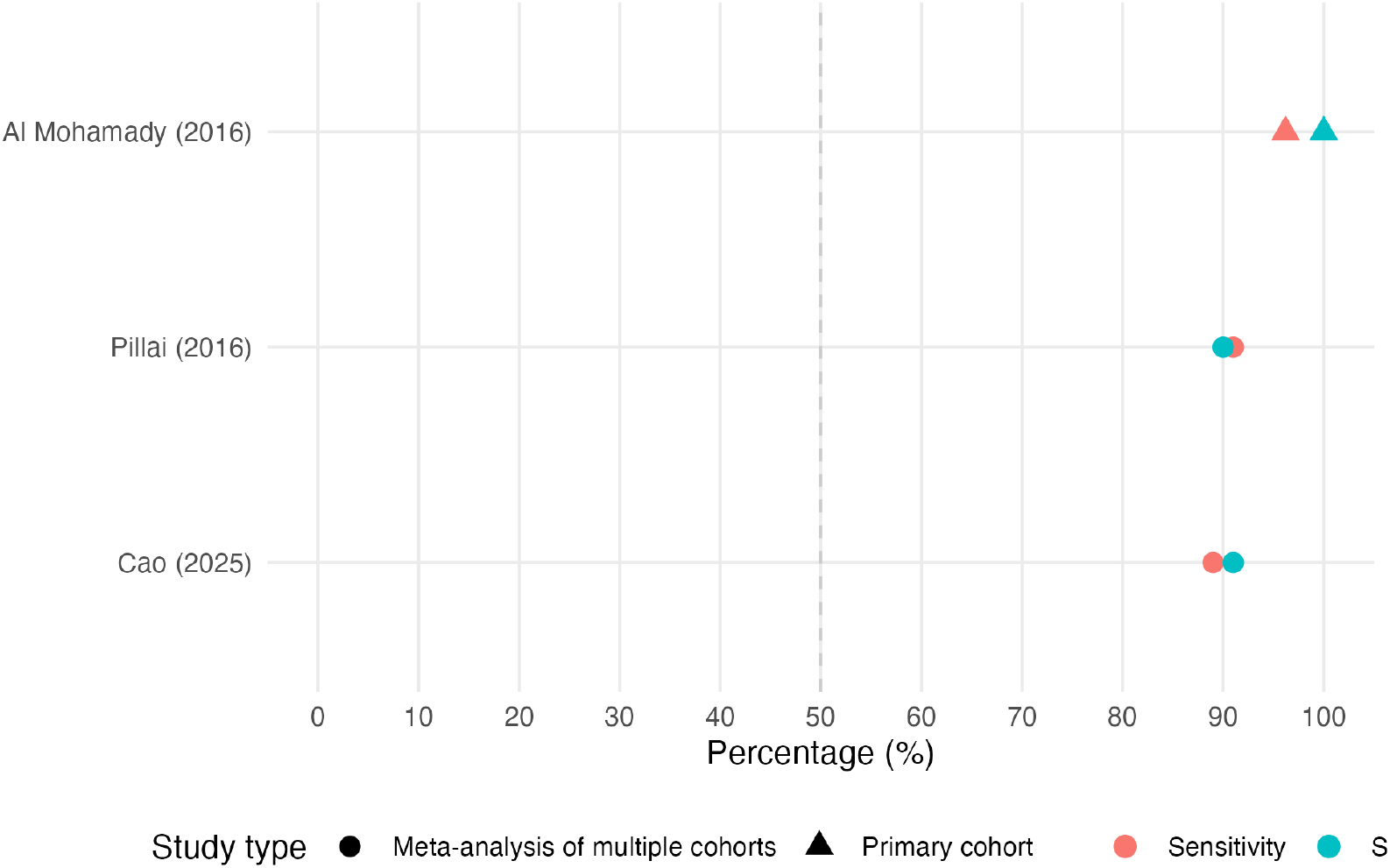
Sensitivity and specificity of serum CA-125.

#### 3.3.2 Serum CA-125

Three studies (two meta-analyses and one primary cohort; total n = 2,529 women) evaluated serum CA-125 as a prognostic biomarker (Table 3B). In a recent meta-analysis of 1,166 women with threatened miscarriage, Cao et al. (2025) reported pooled sensitivity 89.0%, specificity 91.0%, and AUC 0.95 across a range of thresholds (Cao et al., 2025). An earlier meta-analysis by Pillai et al. (2016; n = 1,263) produced similar summary estimates (sensitivity 91.0%, specificity 90.0%), supporting consistency of effect across datasets and analytic methods (Pillai et al., 2016).

In the smaller primary cohort by Al Mohamady et al. (2016; n = 100), a cut-off of 31.2 IU/mL achieved sensitivity 96.2% and specificity 100%. Although this single-centre result should be interpreted cautiously due to sample size, it is aligned with the meta-analytic finding that markedly elevated CA-125 levels are strongly associated with subsequent pregnancy loss among women presenting with threatened miscarriage.

The corresponding graphical summary of sensitivity and specificity estimates for CA-125 across studies is given in Figure 3B. Sensitivity and specificity of serum CA-125 (Appendix).

#### 3.3.3 Comparative interpretation of biochemical markers

When considered together, progesterone and CA-125 show complementary prognostic patterns. Progesterone concentrations function largely as an indicator of impaired luteal or placental support: very low levels are associated with a high probability of miscarriage, but a proportion of losses occur at values above typical cut-offs, which constrains overall sensitivity in pooled analyses. CA-125 appears to act more as a marker of tissue disruption or inflammatory response, with several independent analyses showing sensitivity and specificity around 90% and AUC approaching 0.95. On the basis of these data, CA-125 emerges as a promising single biochemical marker for risk stratification, with progesterone providing additional information at extreme low concentrations or as part of multivariable models, rather than as a stand-alone triage test.

### 3.4. Ultrasound predictors and performance

#### 3.4.1 Ultrasound features across studies

Ultrasound parameters were widely evaluated but heterogeneous in their reporting (Table S4, Appendix). Crown–rump length (CRL) and fetal heart rate (FHR) were the most consistently assessed, with some cohorts additionally reporting gestational sac size or morphology and a smaller number describing subchorionic haematoma and yolk-sac features. In most primary cohorts these parameters were measured at the index visit for threatened miscarriage; in meta-analytic work, they were abstracted from emergency-department cohorts with varying gestational ages at presentation.

Across studies, ultrasound markers were typically examined in combination with clinical or biochemical predictors rather than as isolated tests, reflecting current practice where ultrasound contributes to an integrated assessment of threatened miscarriage. The pattern of which specific ultrasound features were evaluated in each study is summarised graphically in Figure 4C. Ultrasound features evaluated across included studies (Appendix).

**Figure 4C.**
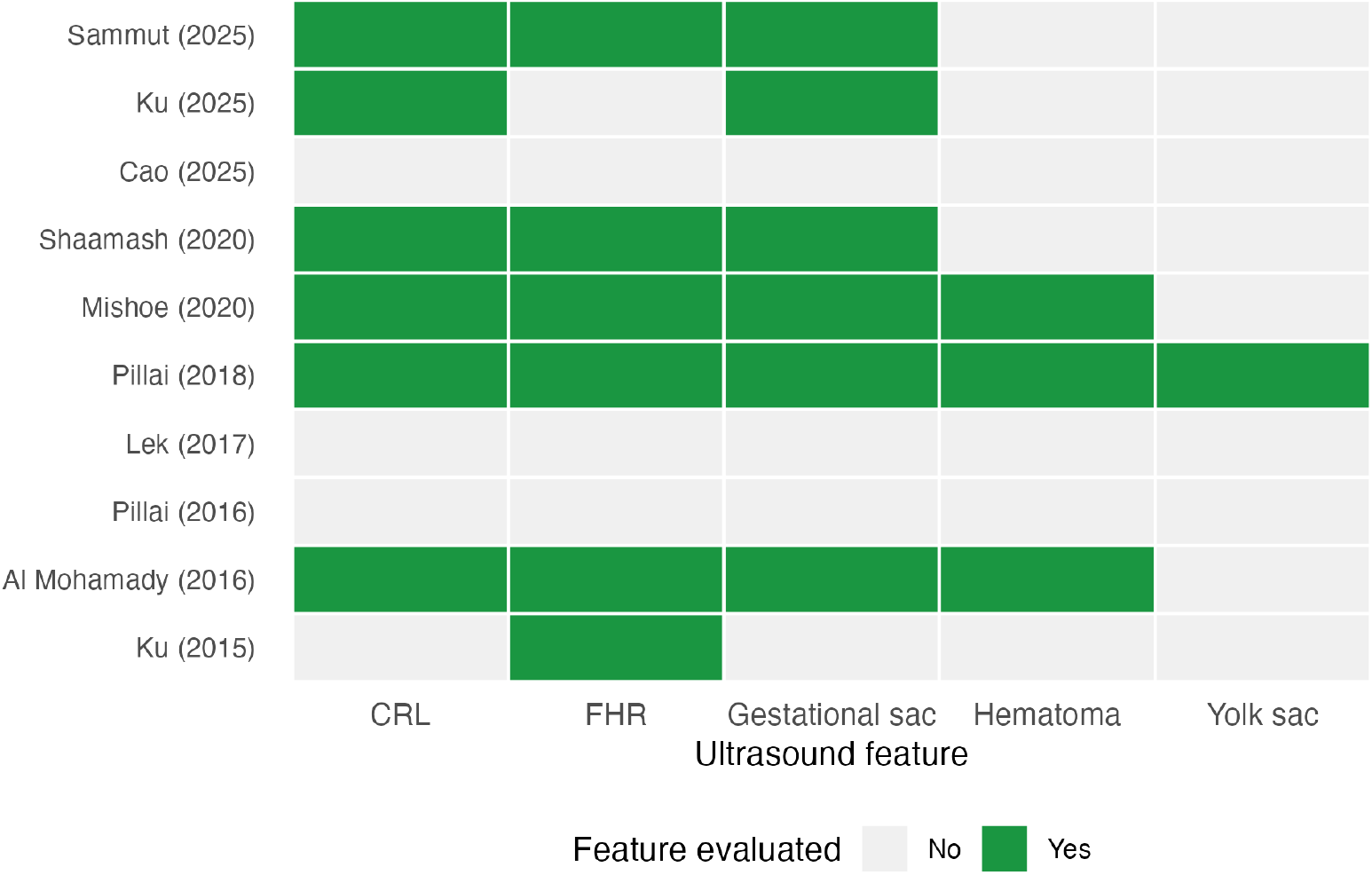
Ultrasound features evaluated across included studies.

#### 3.4.2 Fetal heart rate

Quantitative prognostic data for FHR were available from both primary cohorts and meta-analyses (Table 4). Across studies, low FHR thresholds in the first trimester—most commonly in the range of 100–120 beats per minute—were consistently associated with increased risk of subsequent miscarriage. At these thresholds, sensitivity estimates were generally modest, indicating that some pregnancies that later miscarried had FHR values above the cut-off. Specificity, however, was high, frequently in the upper 80–90% range or higher, and reported AUCs indicated fair-to-good discrimination (Pillai et al., 2016). These findings suggest that a distinctly slow FHR is a specific warning sign for pregnancy loss but does not, by itself, provide a comprehensive risk assessment for all women with threatened miscarriage.

**Table 4A.**
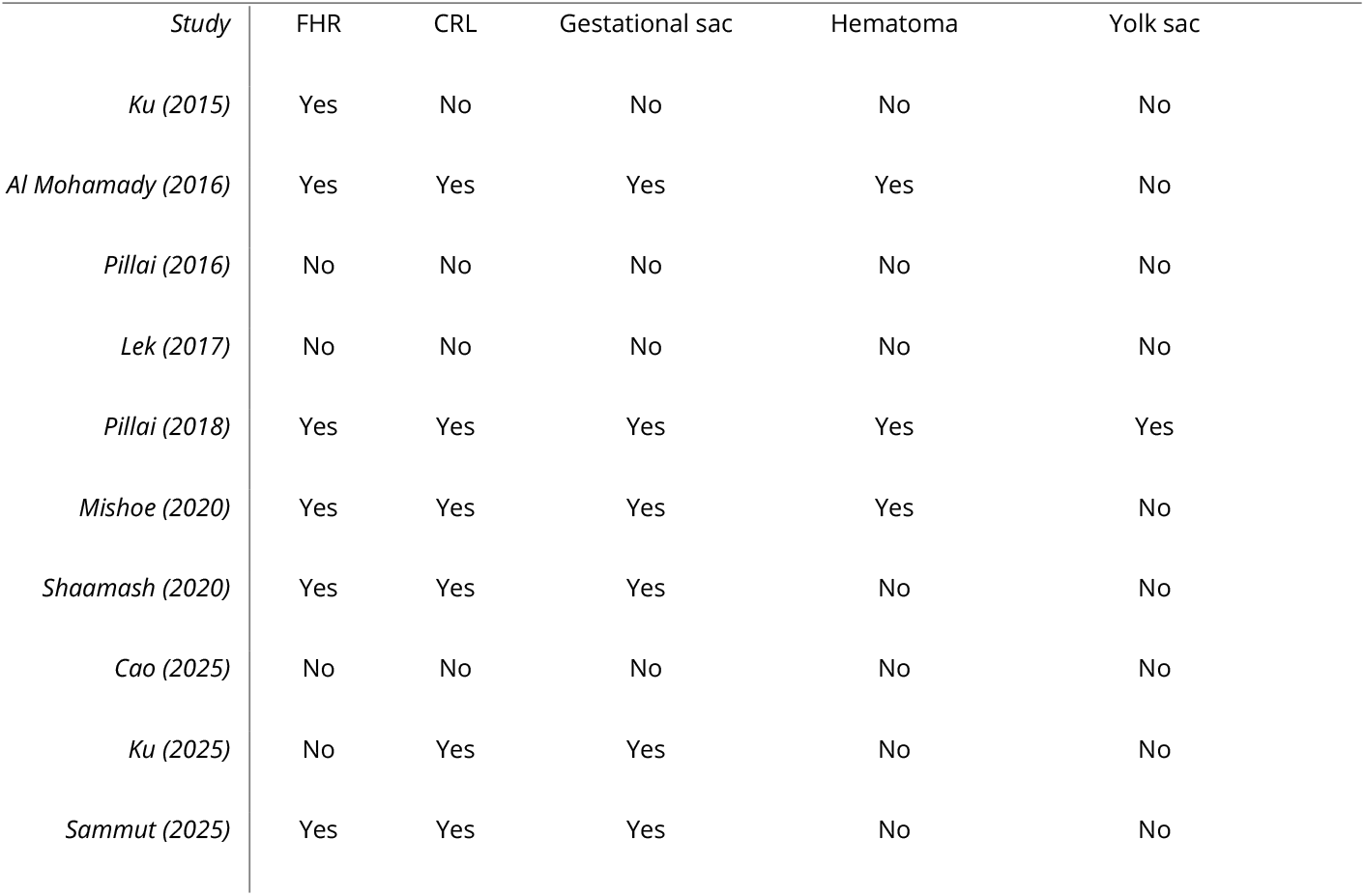
Ultrasound features evaluated across included studies. Table 4. Prognostic performance of fetal heart rate and embryo/sac size parameters.

#### 3.4.3 CRL, gestational sac size, and related metrics

Embryo size and sac-related metrics (CRL, mean gestational sac diameter, and sac–CRL discordance) showed a broadly similar pattern (Table 4). Studies consistently reported that smaller-than-expected CRL or gestational sac dimensions, as well as marked discordance between sac and embryo size, were associated with increased miscarriage risk. As with FHR, these markers typically had high specificity (often ≥85–90%) but only moderate sensitivity. AUC values again fell in the fair-to-good range, and pooled estimates from evidence-synthesis studies were consistent with individual cohort findings. These data indicate that markedly abnormal growth parameters substantially increase the probability of pregnancy loss, but normal or near-normal values do not reliably exclude it.

#### 3.4.4 Summary of ultrasound findings

Taken together, the ultrasound literature indicates that first-trimester FHR and early embryo/sac size measurements provide specific prognostic information for women with threatened miscarriage. Markedly abnormal values identify a subgroup at clearly elevated risk of miscarriage, but the limited sensitivity and between-study variation in thresholds and scan timing mean that ultrasound markers alone are not sufficient as stand-alone prognostic tools. These findings are consistent with an approach in which ultrasound is combined with biochemical and clinical predictors to provide more comprehensive risk stratification.

### 3.5. Multivariable and machine-learning prediction models

Only two contemporary cohorts—Ku et al. and Sammut et al.—developed formal prediction models for miscarriage specifically among women presenting with threatened miscarriage (Table 5). Ku and colleagues (2015) constructed five multivariable logistic regression models using combinations of maternal characteristics, serum progesterone, and early ultrasound findings. Their final “holistic” model, which informed a simple clinical risk score, incorporated maternal age ≥40 years, progesterone <35 nmol/L, gestational age <6.36 weeks, and absent fetal heart activity. In internal validation, this model achieved an AUC of approximately 0.90, with sensitivity and specificity both ≥80% at the prespecified risk threshold (Ku et al., 2015). Across Ku’s model series, discrimination ranged from modest for purely clinical models (AUC ≈0.57) to strong once biochemical and ultrasound predictors were added (AUC 0.80–0.90), supporting the importance of combining domains. The AUCs of each of Ku’s logistic models and Sammut’s models are plotted in Figure 5A. Discriminative performance (AUC) of multivariable models (Appendix).

**Table 5A.**
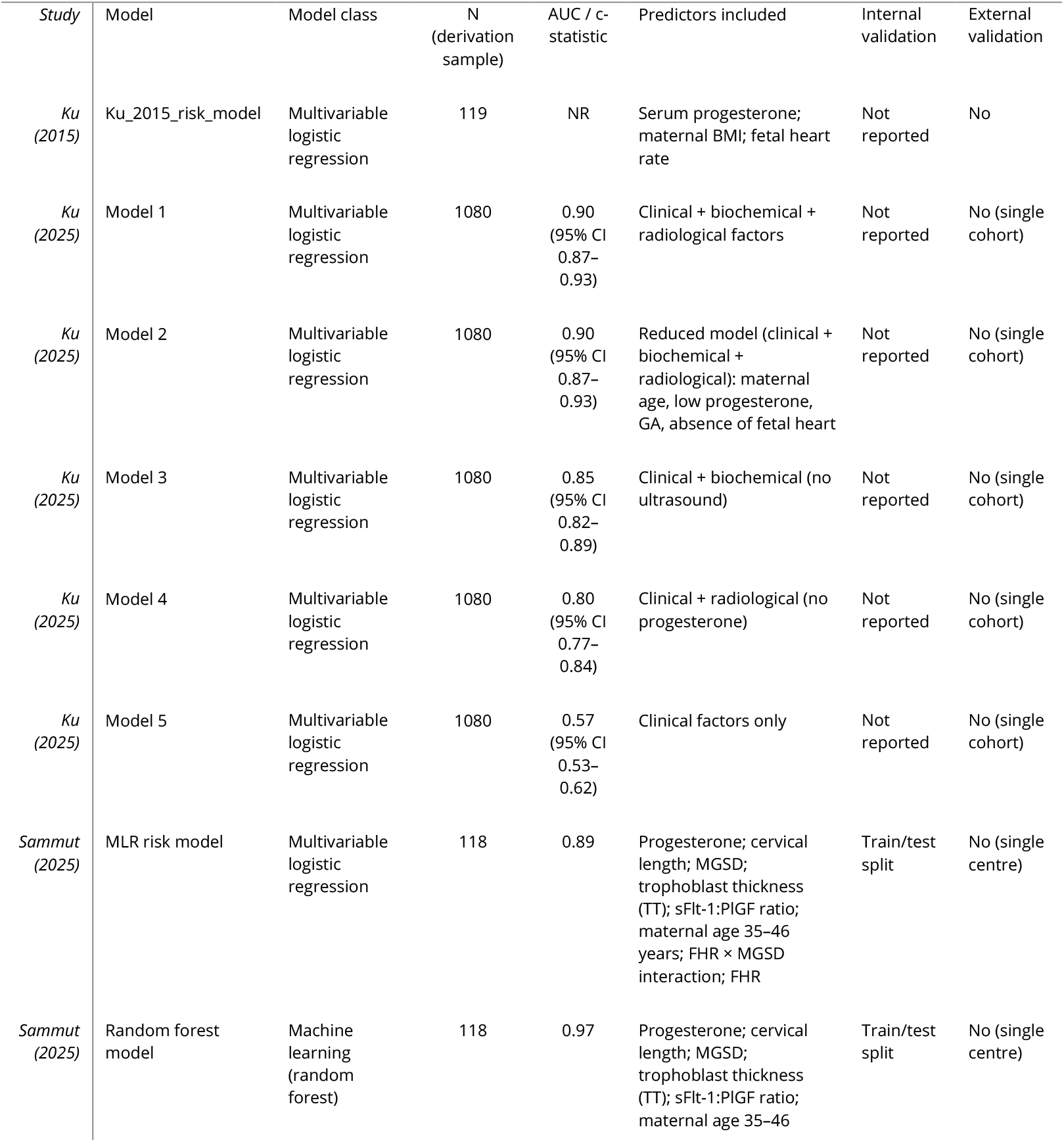
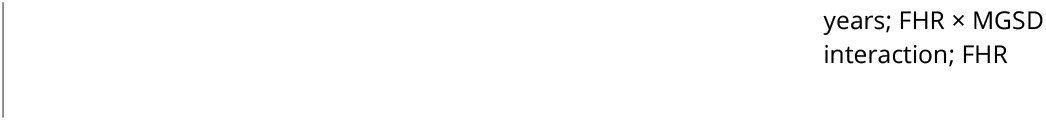
Multivariable and machine-learning models predicting miscarriage after threatened miscarriage. Table 5. Multivariable and machine-learning prediction models, predictor sets, and discrimination.

| Study | Model | Model class | N<br>(derivation<br>sample) | AUC / c-statistic | Predictors included | Internal<br>validation | External<br>validation |
| --- | --- | --- | --- | --- | --- | --- | --- |
| *Ku*<br>(2015) | Ku_2015_risk_model | Multivariable<br>logistic<br>regression | 119 | NR | Serum progesterone;<br>maternal BMI; fetal heart<br>rate | Not<br>reported | No |
| *Ku*<br>(2025) | Model 1 | Multivariable<br>logistic<br>regression | 1080 | 0.90<br>(95% CI<br>0.87–<br>0.93) | Clinical + biochemical +<br>radiological factors | Not<br>reported | No (single<br>cohort) |
| *Ku*<br>(2025) | Model 2 | Multivariable<br>logistic<br>regression | 1080 | 0.90<br>(95% CI<br>0.87–<br>0.93) | Reduced model (clinical +<br>biochemical +<br>radiological); maternal<br>age, low progesterone,<br>GA, absence of fetal heart | Not<br>reported | No (single<br>cohort) |
| *Ku*<br>(2025) | Model 3 | Multivariable<br>logistic<br>regression | 1080 | 0.85<br>(95% CI<br>0.82–<br>0.89) | Clinical + biochemical (no<br>ultrasound) | Not<br>reported | No (single<br>cohort) |
| *Ku*<br>(2025) | Model 4 | Multivariable<br>logistic<br>regression | 1080 | 0.80<br>(95% CI<br>0.77–<br>0.84) | Clinical + radiological (no<br>progesterone) | Not<br>reported | No (single<br>cohort) |
| *Ku*<br>(2025) | Model 5 | Multivariable<br>logistic<br>regression | 1080 | 0.57<br>(95% CI<br>0.53–<br>0.62) | Clinical factors only | Not<br>reported | No (single<br>cohort) |
| *Sammut*<br>(2025) | MLR risk model | Multivariable<br>logistic<br>regression | 118 | 0.89 | Progesterone; cervical<br>length; MGSD;<br>trophoblast thickness<br>(TT); sFlt-1:PIGF ratio;<br>maternal age 35–46<br>years; FHR × MGSD<br>interaction; FHR | Train/test<br>split | No (single<br>centre) |
| *Sammut*<br>(2025) | Random forest<br>model | Machine<br>learning<br>(random<br>forest) | 118 | 0.97 | Progesterone; cervical<br>length; MGSD;<br>trophoblast thickness<br>(TT); sFlt-1:PIGF ratio;<br>maternal age 35–46 | Train/test<br>split | No (single<br>centre) |

**Figure 5A.**
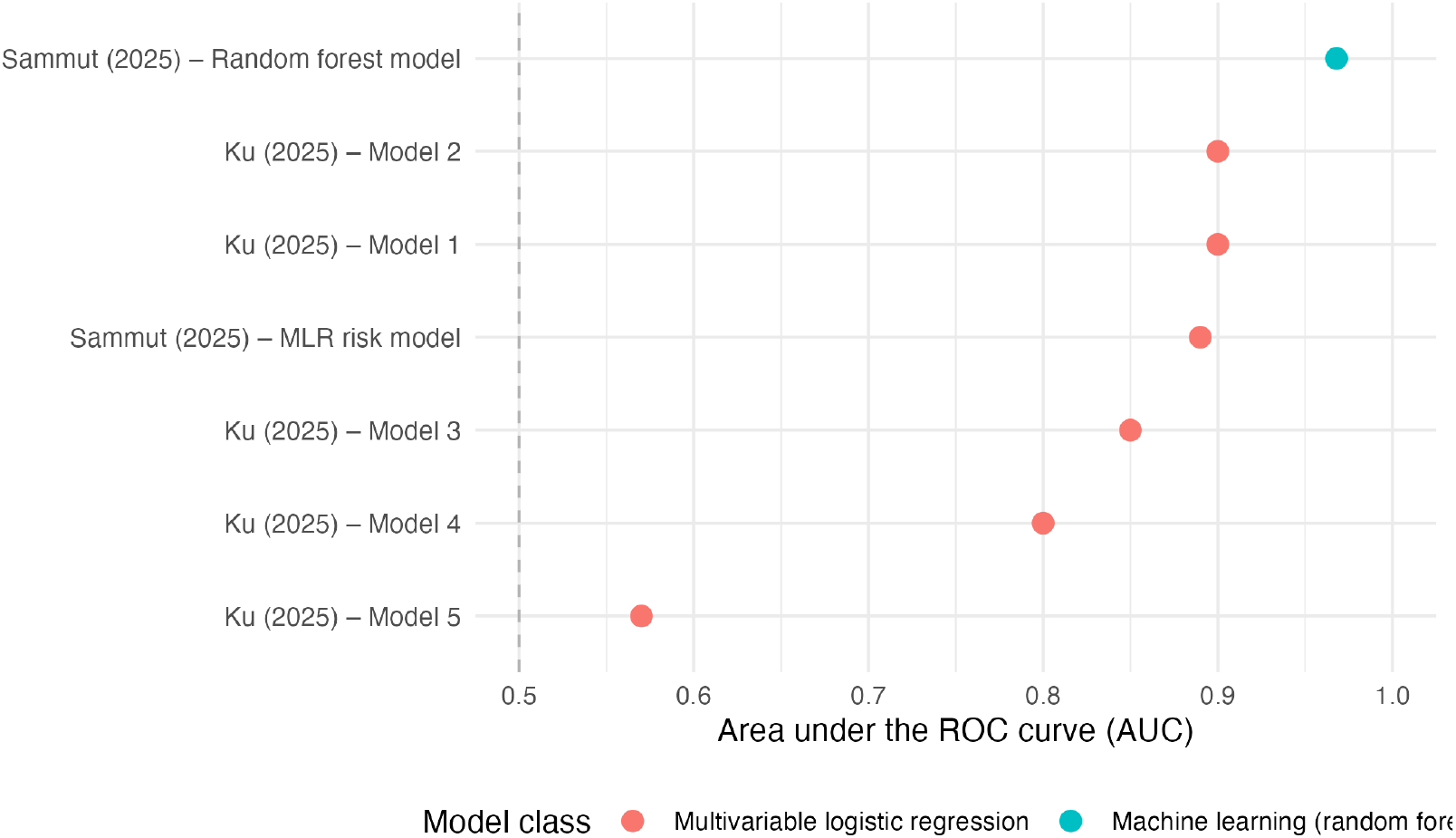
Discriminative performance (AUC) of multivariable models.

**Figure 5B.**
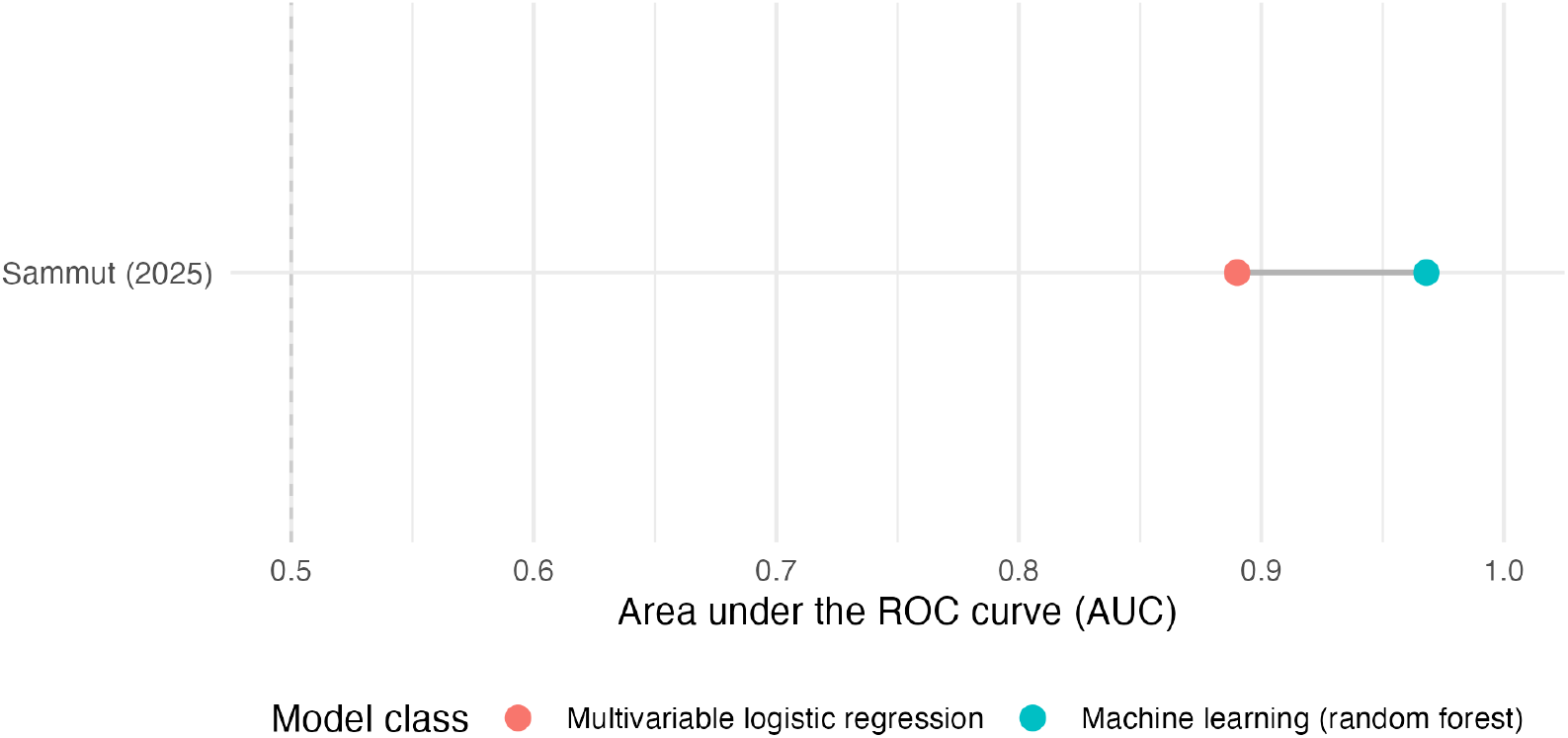
Logistic regression vs random forest (Sammut et al.).

**Figure 5C.**
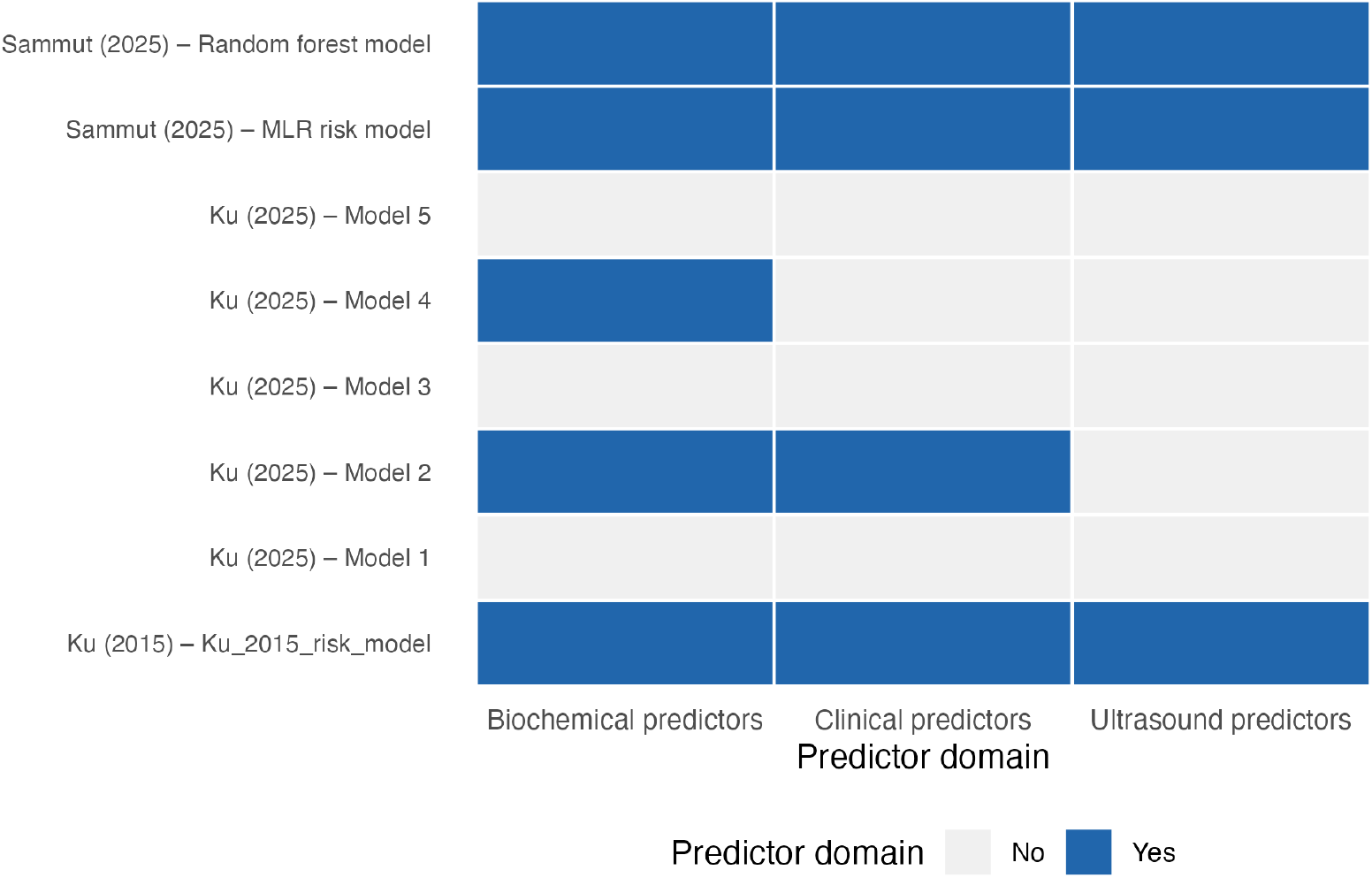
Predictor domains in multivariable models.

Sammut et al. (2025) extended this work by directly comparing a multivariable logistic regression model with a machine-learning random-forest classifier in a smaller but extensively characterised first-trimester cohort that included clinical, biochemical, angiogenic, and detailed ultrasound markers. Using identical predictor sets and a stratified train–test split, the logistic model achieved a test-set AUC of approximately 0.89, while the random-forest model achieved an AUC of about 0.97, with higher overall accuracy, higher positive predictive value, and very high negative predictive value on the held-out test data (Sammut et al., 2025). The head-to-head comparison of these two models is shown in Figure 5B. Logistic regression vs random forest (Sammut et al.) (Appendix).

Across these multivariable studies, a consistent pattern emerges. Models restricted to clinical history variables alone show limited discrimination. Models that integrate at least one robust biochemical marker with early transvaginal ultrasound features achieve AUC values above 0.80, indicating good prognostic performance. More complex machine-learning models may achieve higher apparent discrimination in internal validation. However, all models were developed in single-centre cohorts, none have undergone external validation, and the most complex algorithms were trained on relatively small samples. These characteristics suggest that the reported performance—particularly for the random-forest model—should be interpreted as preliminary and hypothesis-generating, rather than as directly applicable tools for routine clinical practice. The composition of predictor domains (clinical, biochemical, and ultrasound) within each multivariable or machine-learning model is summarised in Figure 5C. Predictor domains in multivariable models (Appendix).

### 3.6. Risk of bias, applicability, and publication bias

Four studies reported structured quality assessment; three used QUADAS-2 and one used the QUAPAS tool, consistent with contemporary standards for prognostic accuracy reviews (Table S5, Appendix). Figure 6A. Risk-of-bias tools used in included studies (Appendix) shows the frequency with which each tool was applied. Overall risk of bias was generally judged as low to unclear for the recent multivariable models and ultrasound cohorts, but concerns were concentrated in patient selection, conduct and interpretation of the index test and reference standard, and flow/timing. In particular, the dataset reported by Pillai et al. (2016) was rated as having concerns in all four QUADAS-2 domains because of non-consecutive recruitment and limited reporting of follow-up and attrition. These issues indicate that some of the more favourable single-biomarker estimates should be interpreted cautiously and support conservative grading of the certainty of evidence; the specific domains affected are depicted in Figure 6B. Bias domains with explicitly described concerns (Appendix).

**Figure 6A.**
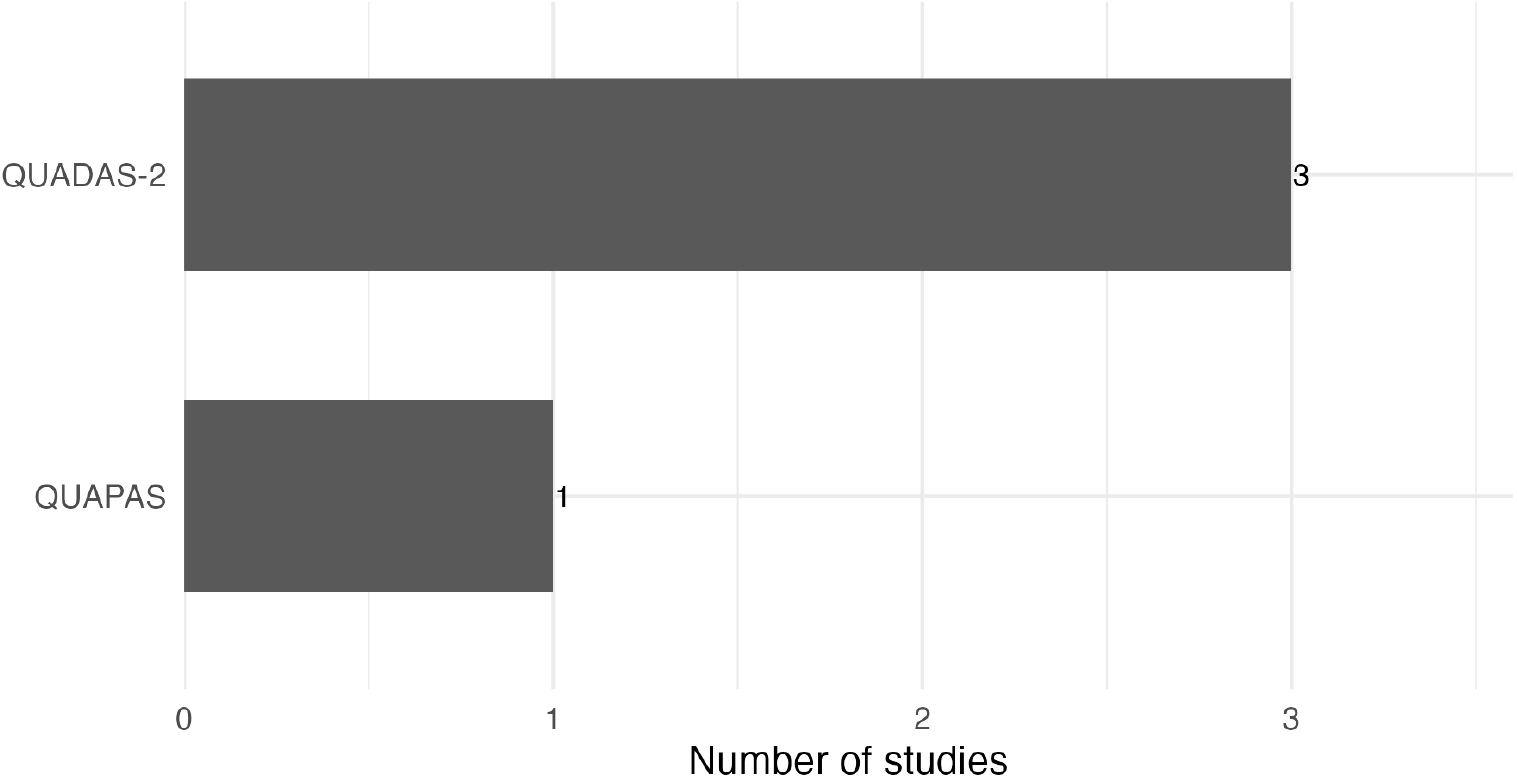
Risk-of-bias tools used in included studies.

Formal assessment of small-study and publication bias was feasible only for the largest CA-125 meta-analysis, in which Deeks’ funnel plot appeared symmetrical (P = 0.81), providing no strong evidence of publication bias for that marker (Table S6, Appendix). For serum progesterone, ultrasound features, and multivariable models, the limited number of contributing cohorts precluded reliable funnel-plot or regression-based tests, in line with guidance that such methods are underpowered when few studies are available.

Overall, the certainty of evidence for prognostic performance of individual markers and models in threatened miscarriage is most affected by risk of bias, inconsistency between studies, and imprecision, rather than by demonstrable publication bias.

## Discussion

In this systematic review, we found that prognostic research in first-trimester threatened miscarriage is both quantitatively modest and methodologically heterogeneous, despite the high absolute risk faced by affected women. Across six primary cohorts, miscarriage risks consistently clustered around 15–25% among women presenting with first-trimester bleeding and a viable intrauterine pregnancy, confirming that threatened miscarriage is a genuinely high-risk state rather than a benign variant of normal pregnancy. In contemporary prospective cohorts of women with first-trimester bleeding and a viable intrauterine pregnancy, roughly one in five pregnancies ends in loss despite initial sonographic viability. This elevated baseline risk provides the critical context in which single biomarkers, ultrasound features, and multivariable models must demonstrate added prognostic value.

Biochemical markers showed clear but incomplete prognostic utility. Across three studies (one meta-analysis and two primary cohorts; n=1,709), low serum progesterone levels were consistently associated with miscarriage, but performance depended heavily on the threshold used. The Pillai meta-analysis (n=1,263) reported pooled sensitivity of only 30% with specificity 86% at commonly used cut-offs, indicating that most losses occur above traditional progesterone thresholds even though very low values are strongly predictive of non-viability. By contrast, single-centre cohorts reported strikingly high rule-in performance at more extreme cut-offs: Al-Mohamady et al. (n=100) observed sensitivity 97.5% and specificity 100% at 11.5 ng/mL, while Lek et al. (n=346) validated a 35 nmol/L threshold with sensitivity 65.7%, specificity 92.0%, and AUC 0.83. These data suggest that progesterone behaves as a “floor” marker: very low concentrations confer near-certain risk of loss, but many miscarriages occur despite progesterone values in or near the reference range.

CA-125 showed even more impressive discriminative performance, albeit on a similarly limited evidence base. Across three studies (two meta-analyses and one primary cohort; n=2,529), pooled sensitivity and specificity were consistently about 90% for miscarriage prediction. Cao et al. (2025; n=1,166) reported sensitivity 89%, specificity 91%, and AUC 0.95, while the earlier Pillai meta-analysis (n=1,263) found nearly identical summary estimates (sensitivity 91%, specificity 90%). In the Al-Mohamady cohort (n=100), a threshold of 31.2 IU/mL yielded sensitivity 96.2% and specificity 100%, reinforcing the signal that markedly elevated CA-125 is almost pathognomonic for impending pregnancy failure in this context. Across more than 2,500 women with threatened miscarriage, CA-125 consistently achieved sensitivities and specificities around 90% with AUCs approaching 0.95, indicating that it behaves as a near-single-test prognostic marker in this setting. However, these results arise from overlapping cohorts and lack extensive external validation, so they remain hypothesis-generating rather than practice-changing.

Ultrasound parameters were near-universal across cohorts and meta-analytic datasets, and they displayed a characteristic pattern of high specificity but modest sensitivity. Fetal heart rate and crown–rump length were the most consistently assessed predictors, with several studies also evaluating gestational sac size, sac–embryo discordance, yolk-sac morphology, and subchorionic hematoma. Abnormally slow fetal heart rate in early gestation (typically ≤110–120 bpm) and smaller-than-expected CRL or gestational sac diameter were strongly associated with subsequent loss, with specificities frequently ≥85–90% and AUC values in the fair-to-good range. Normal measurements, however, did not reliably exclude miscarriage, reflecting the multifactorial nature of early pregnancy failure. Taken together, first-trimester ultrasound viability parameters function as powerful rule-in markers—especially when fetal heart rate and embryo/sac size are clearly abnormal—but they cannot safely be used as stand-alone rule-out tests for women with threatened miscarriage.

Only a small number of studies moved beyond single markers to develop formal prediction models. Ku and colleagues showed that purely clinical models (age, bleeding pattern, prior miscarriage) yielded poor discrimination (AUC ≈0.57), whereas adding progesterone and basic ultrasound parameters increased AUCs into the 0.80–0.90 range, with the best “holistic” model achieving an internally validated AUC around 0.90 and balanced sensitivity and specificity ≥80%. Sammut et al. extended this approach by comparing logistic regression with a random-forest classifier in a deeply phenotyped but relatively small cohort. Using the same multimodal predictor set (clinical, biochemical, angiogenic and ultrasound markers), the logistic model achieved good discrimination (test-set AUC ≈0.89), while the random forest improved apparent performance to an AUC ≈0.97 with a negative predictive value close to 1.00. Across multivariable models, we observed a clear performance gradient: clinical-only models performed poorly (AUC ≈0.57), integrated clinical–biochemical– ultrasound models consistently achieved AUCs >0.80, and the most complex machine-learning model approached an AUC of 0.97 on internal validation. These figures underline the theoretical value of multimodal, non-linear modelling for threatened miscarriage, but they also highlight the danger of overfitting in small, single-centre datasets.

Risk-of-bias assessments explain much of the residual uncertainty. Several older cohorts and some meta-analytic datasets were at high or unclear risk of bias across QUADAS-2 domains, particularly in patient selection (non-consecutive recruitment, restricted populations), index-test conduct and interpretation (post-hoc cut-off selection, lack of blinding), and flow/timing (incomplete follow-up, poorly defined outcome windows). Prediction-model studies frequently dichotomised continuous predictors, used events-per-parameter ratios that were too low, and provided limited information on calibration, handling of missing data, and internal validation. These methodological limitations restrict both the credibility of headline AUC values and the feasibility of robust meta-analysis or model aggregation. In keeping with modern prognostic-research standards, the overall certainty of evidence for most markers and models is therefore downgraded not for lack of signal, but for risk of bias, inconsistency, and imprecision.

The clinical implications are pragmatic rather than revolutionary. Our synthesis indicates that clinicians already hold a powerful set of routinely available prognostic signals—fetal heart rate, crown–rump length, gestational sac morphology, and basic biochemical markers such as progesterone and CA-125—but these signals are rarely combined and almost never deployed within validated, user-friendly prediction tools. A woman with markedly low progesterone, very high CA-125, and clearly abnormal ultrasound findings has a substantially higher absolute risk of miscarriage than a woman with normal values across all domains, and this risk can be communicated quantitatively; however, normal or borderline results do not guarantee a favourable outcome. CA-125, in particular, appears poised to play a central prognostic role if future work confirms its ≈90% sensitivity and specificity across more diverse populations. At the same time, the striking AUCs seen in single-centre machine-learning models must not be interpreted as clinically ready until they are externally validated, calibrated, and tested for impact.

Looking forward, the research agenda is clear. Large, prospectively registered, multicentre cohorts are needed, with harmonised definitions of threatened miscarriage, standardised measurement of a prespecified multimodal predictor set, and analytic plans that respect the continuous nature of biochemical and ultrasound variables. Modern regression and machine-learning techniques should be used with transparent penalisation, internal validation, and full reporting of discrimination, calibration, and decision-curve–based clinical utility. The field now needs fewer new biomarkers and many more rigorously designed, multicentre validation studies that test whether integrated risk models actually improve counselling, triage, and outcomes for women with threatened miscarriage. Only when such evidence is available will it be possible to move from proof-of-concept models to guideline-endorsed tools that deliver robust, honest, and clinically useful risk estimates to the millions of women worldwide who experience first-trimester bleeding each year.

## Conclusion

In this systematic review of ten cohorts and evidence-synthesis studies of first-trimester threatened miscarriage, we found that miscarriage risks consistently clustered around 15– 25% among women presenting with vaginal bleeding and an initially viable intrauterine pregnancy, confirming that threatened miscarriage is a genuinely high-risk state rather than a benign variant of normal pregnancy. Across >4,000 women, biochemical and ultrasound predictors provided substantial prognostic information but fell short of a definitive stand-alone solution. CA-125 emerged as the most powerful single biochemical marker, with pooled sensitivities and specificities both around 90% and AUCs approaching 0.95, whereas progesterone and early ultrasound parameters (fetal heart rate, crown–rump length, gestational sac morphology) functioned primarily as highly specific “rule-in” signals rather than reliable “rule-out” tests.

Multivariable and machine-learning models that combined clinical, biochemical, and ultrasound domains consistently outperformed single-marker strategies. Clinical-only models performed poorly (AUC ≈0.57), whereas integrated models routinely achieved AUCs above 0.80, and the most complex random-forest implementation approached an AUC of 0.97 on internal validation, albeit in small, single-centre datasets. Yet none of these models has been externally validated across diverse health-system contexts, few reported robust calibration, and no impact studies have shown that their use improves patient-centred outcomes. At present, therefore, no single biomarker, ultrasound feature, or prediction model meets the evidentiary standard required for guideline-endorsed implementation in routine care.

Clinically, our findings support using existing ultrasound and biochemical data to provide nuanced, probabilistic counselling—especially for women with clearly abnormal parameters—while avoiding over-reliance on any single test to “rule out” miscarriage. Strategically, they define the next step for the field: the priority is no longer to discover yet another biomarker, but to embed a prespecified multimodal predictor set in large, rigorously designed, multicentre cohorts and to subject integrated risk models to transparent validation, calibration, and impact evaluation. Delivering such models will be essential if we are to move from fragmented prognostic signals to trustworthy, evidence-based risk tools that can be offered, with honesty and precision, to the millions of women worldwide who experience threatened miscarriage each year.

Abbreviations: TM (threatened miscarriage), ED (emergency department), EPU (early pregnancy unit), hCG (human chorionic gonadotrophin), FHR (fetal heart rate), CRL (crown– rump length), AUC (area under the receiver operating characteristic curve), ROC (receiver operating characteristic), CA-125 (cancer antigen 125), ML (machine learning), RF (random forest), PPV (positive predictive value), NPV (negative predictive value), CI (confidence interval), QUADAS-2 (Quality Assessment of Diagnostic Accuracy Studies-2), QUAPAS (Quality Assessment of Prognostic Accuracy Studies).

## Data Availability

All data referred to in this manuscript, including the anonymised extracted dataset, extraction template, and full analysis code, are openly available via the OSF registry and protocol (https://doi.org/10.17605/OSF.IO/45V63
), the companion GitHub repository (https://github.com/drsunday-ade/threatened-miscarriage-prognostic-review/
), and the archived Zenodo record (https://doi.org/10.5281/zenodo.17645695
).

https://github.com/drsunday-ade/threatened-miscarriage-prognostic-review

https://doi.org/10.5281/zenodo.17645695

## Contributors

SAA conceived the review question, designed the protocol, and is the guarantor of the work. SAA and DN developed the search strategy, eligibility criteria, and data extraction framework. SAA conducted the literature searches, screened titles/abstracts and full texts, and completed the primary data extraction and risk-of-bias assessments; DN independently verified study eligibility, data extraction, and quality assessments, with disagreements resolved by consensus. SAA wrote the statistical analysis plan, implemented all analyses, and produced the figures and tables. SAA drafted the manuscript with critical revisions from DN for intellectual content. Both authors had full access to all the data, approved the final version of the manuscript, and agree to be accountable for all aspects of the work.

## Data sharing and code availability

All study-level extracted data and the complete R code used to reproduce the analyses, figures, and tables are openly available:

- OSF registry and protocol: https://doi.org/10.17605/OSF.IO/45V63
- GitHub repository (data code, extraction template, and outputs): https://github.com/drsunday-ade/threatened-miscarriage-prognostic-review/
- Zenodo repository for data: https://doi.org/10.5281/zenodo.17645695/

The dataset contained in the repository is derived entirely from published articles and does not include any individual patient identifiers. Researchers are free to reuse the materials with appropriate citation.

## Ethics approval

This project is a systematic review and meta-analysis of previously published studies and does not involve collection of new individual-level data. In accordance with Oregon State University and international guidance, formal ethics committee approval and informed consent were not required.

## Patient and public involvement

No patients or members of the public were involved in the design, conduct, reporting, or dissemination plans of this research, because the study used only published aggregate data. Future work will prioritise co-design of communication materials with patient representatives from early pregnancy and miscarriage support groups.

## Acknowledgments

The authors thank colleagues in the Department of Epidemiology, College of Health, Oregon State University, for methodological feedback during protocol development and interpretation of findings. We also acknowledge the authors of the primary studies included in this review, whose work made this synthesis possible.

## Appendix: Supplementary figures and tables

### Supplementary figures

Supplementary tables(check the repositories for the additional figures/tables: https://github.com/drsunday-ade/threatened-miscarriage-prognostic-review)

Table S1. Reasons for exclusion of 52 full-text articles at screening, grouped by population, design, data availability, and overlap with included cohorts.

Table S2. Detailed design and setting of primary cohorts and evidence-synthesis studies, including recruitment period, care setting (emergency department, early pregnancy unit, outpatient clinic), and operational definitions of threatened miscarriage.

Table S3. Characteristics of evidence-synthesis and meta-analytic studies, including included cohorts, target population, primary outcomes, and summary accuracy metrics.

Table S4. Distribution, measurement methods, and operational definitions of ultrasound predictors across included studies, including fetal heart rate, embryo and sac size parameters, yolk-sac features, and subchorionic haematoma.

Table S5. Risk-of-bias and applicability assessments across QUADAS-2 and QUAPAS domains for individual studies, with domain-level judgements and overall ratings.

Table S6. Publication-bias assessment for CA-125 and other markers, including Deeks’ funnel-plot asymmetry test and regression-based statistics where applicable.

